# Are time series foundation models the new frontier for operational epidemic forecasting?

**DOI:** 10.64898/2026.11.24.26368762

**Authors:** Stefania Fiandrino, Ana Parovic, Alessandra Urbinati, Sara Venturini, Rebecca K. Borchering, Matthew Biggerstaff, Nicolò Gozzi, Alessandro Vespignani

## Abstract

Time series foundation models (TSFMs) have emerged as a promising frontier in forecasting, particularly for their strong zero-shot performance (i.e., the ability to forecast without task-specific training) and domain generalization. Despite a few early studies, their potential for operational epidemic forecasting remains an open question. Here, we prospectively evaluate six state-of-the-art TSFMs (Chronos-2, Lag-Llama, Moirai-2, TabPFN-TS, TimeGPT, and TimesFM) for real-time short-term epidemic forecasting, providing each model with recent surveillance data. We benchmark their performance against that of individual and ensemble models submitted to FluSight and CovidHub, two U.S. CDC-coordinated collaborative hubs covering the U.S. national level and 52 U.S. jurisdictions. Forecasts were generated prospectively during the 2025/26 winter season for weekly laboratory-confirmed influenza or COVID-19 hospital admissions. Additionally, we conducted a retrospective evaluation of the 2024/25 season to assess the robustness of our findings. Our results show that TSFMs’ performance is highly heterogeneous across diseases, locations, epidemic phases, and seasons. Among the TSFMs considered, Chronos-2, TimesFM, and TabPFN-TS are the most competitive; TimeGPT exhibits strong point-forecast accuracy but limited calibration; Moirai-2 shows context-dependent performance; and Lag-Llama consistently underperforms across all settings. Although the best-performing TSFMs achieve performance comparable to the multi-model ensemble in some forecasting instances, none does so consistently across diseases, locations, forecasting rounds, and evaluation metrics. The multi-model ensemble remains the most reliable strategy for communicating forecasts up to 3-weeks ahead to public health agencies and to the public. These findings highlight that TSFMs, despite promising performance, are not a single solution for epidemic forecasting, and that rigorous evaluation remains a prerequisite for their integration into operational forecasting efforts.

## 1 Introduction

Accurate and timely epidemic forecasts are fundamental to support public health decision-making, situational awareness, and resource allocation during both emerging and recurrent epidemic threats.^1–8^ Time series foundation models (TSFMs) are large-scale models trained on vast, heterogeneous data to learn general representations, adapted specifically for time series tasks such as forecasting.^9–12^ Recently, TSFMs have attracted growing interest across many domains due to their strong zero-shot forecasting performance (i.e., the ability to reliably forecast without task-specific training), generative capabilities, and potential to integrate multimodal data and transfer knowledge across domains.^9^

In this context, TSFMs represent a promising opportunity for operational epidemic forecasting applications.^13^ Epidemic time series are characterized by a complex temporal structure, including non-stationary dynamics, nonlinearity, changepoints, and substantial variability across diseases, locations, and periods.^13^ TSFMs are specifically designed to generalize across heterogeneous domains without task-specific training, can capture complex temporal patterns, and show strong capability for zero- or few-shot forecasting. Recent studies have explored the application of TSFMs to epidemic forecasting.^14–19^ While some analyses find that TSFMs can generate accurate short-term forecasts and complement existing modeling approaches, others show only marginal improvements over classical statistical methods. These contrasting findings highlight that the potential of TSFMs for epidemic forecasting remains an open question. Most importantly, the performance of multiple TSFMs has not yet been systematically evaluated in applied forecasting tasks against a broad range of alternative approaches for multiple diseases and geographical resolutions. This is a critical gap that leaves their operational value an open question.

In this work, we systematically evaluate the short-term forecasting performance of six state-of-the-art TSFMs (Chronos-2, Lag-Llama, Moirai-2, TabPFN-TS, TimeGPT, TimesFM) in a prospective operational epidemic forecasting analysis. We apply these models using past data as context, without additional fine-tuning. We test two disease targets: influenza and COVID-19 hospitalizations during the 2025/26 winter season for the United States at the national level and for its 50 states, Washington DC, and Puerto Rico. We benchmark TSFMs against the weekly probabilistic projections submitted to two major collaborative forecasting hubs: FluSight, coordinated by the U.S. CDC’s (Centers for Disease Control and Prevention) Influenza Division for influenza,^20^ and the COVID-19 Forecast Hub (CovidHub), coordinated by the CDC’s Center for Forecasting and Outbreak Analytics (CFA).^21^ Collaborative hubs collect projections from multiple independent teams for a common set of forecasting targets. Contributing teams use a wide range of approaches (e.g., statistical, mechanistic, semi-mechanistic, machine learning, deep learning, and hybrid models).^22–33^ Individual model outputs are typically aggregated into a multi-model ensemble, which represents the hub consensus projection. This ensemble has consistently outperformed individual models across diseases, targets, and geographical contexts, making it the reference approach for real-time epidemic forecasting.^26,29,31,32^

Following the approach adopted by FluSight and CovidHub, forecasting performance is evaluated using the weighted interval score (WIS),^34^ the absolute error of the median, and prediction coverage. To further assess the robustness of our findings, we validate the prospective evaluation of the 2025/26 season with a retrospective analysis of the 2024/25 season. Our analysis shows that TSFM performance is highly heterogeneous across models, diseases, locations, and seasons. These findings indicate that TSFMs should not be treated as a monolithic category, but rather as a diverse set of approaches whose effectiveness depends on the specific model and forecasting context. No single TSFM demonstrates the consistency of the multi-model ensemble across diseases, locations, forecasting rounds, and evaluation metrics. While individual TSFMs may outperform the ensemble in specific settings, dates or targets, none does so systematically. For public health agencies, requiring consistent and reliable forecasts, the multi-model ensemble remains the most robust strategy to communicate epidemic forecasts to decision makers. Our work highlights that TSFMs are not a single solution for operational epidemic forecasting but have the potential to expand the forecasting toolkit, and that rigorous evaluation is a prerequisite to identify which models can contribute to collective forecasting efforts.

## 2 Results

In this section, we first describe the forecasting task and the evaluation framework used to benchmark TSFMs in the frameworks provided by FluSight and CovidHub collaborative hubs (Sec. 2.1). We then present the performance of the six TSFMs aggregated across all U.S. locations, comparing them with the hub ensemble, the baseline model, and the individual contributing models (Sec. 2.2). Next, we disaggregate this analysis at the subnational level to characterize model performance across locations (Sec. 2.3). We then evaluate the consistency of forecasting performance across rounds and locations (Sec. 2.4) and stratify performance by epidemic phase to examine whether performance varies in different periods of the epidemic dynamic, i.e., increase or decrease trends (Sec. 2.5). In the main text, we present results for the 2025/26 influenza and COVID-19 seasons. Analogous analyses for the corresponding retrospective evaluations for the 2024/25 season are provided in the Supplementary Information (Sec. S3).

### 2.1 Forecasting task and evaluation framework

The FluSight and the CovidHub forecasting challenges collect short-term forecasts of influenza and COVID-19 hospitalizations from multiple individual models. Participating teams submit probabilistic forecasts of weekly laboratory-confirmed hospital admissions at horizons ranging from 0- to 3-weeks ahead of the reference week (i.e., the week of forecast submission), at both the U.S. national and jurisdiction-specific levels (all 50 states, Washington DC, and Puerto Rico). During the 2025/26 season, a total of 55 models contributed forecasts for influenza hospital admissions to FluSight and 17 models for COVID-19 hospital admissions to the CovidHub. Further details on the collaborative hubs are provided in Sec. 4.2, and information on the participating models is available in the Supplementary Information (Sec. S1).

We conducted a prospective evaluation of six TSFMs during the 2025/26 season under operational real-time forecasting conditions, which required using versioned data available at the time each forecast was made. Weekly forecasts were produced over 27 forecasting rounds, from November 22, 2025, through May 23, 2026, for both hubs. As illustrated in Figure 1, each week, new surveillance data were released publicly through the CDC’s Weekly Hospital Respiratory Data (HRD) from the National Healthcare Safety Network (NHSN),^35^ prompting a new round of forecast submission according to each hub’s task specifications (i.e., required targets, locations, forecasting horizons, and submission format). Officially contributing models submitted their forecasts to a dedicated public GitHub repository for both FluSight^36^ and CovidHub.^37^ To align our evaluation with this operational framework and ensure a fair prospective comparison, we followed the same submission guidelines in a separate private GitHub repository that collected TSFM forecasts during the season.

**Figure 1.**
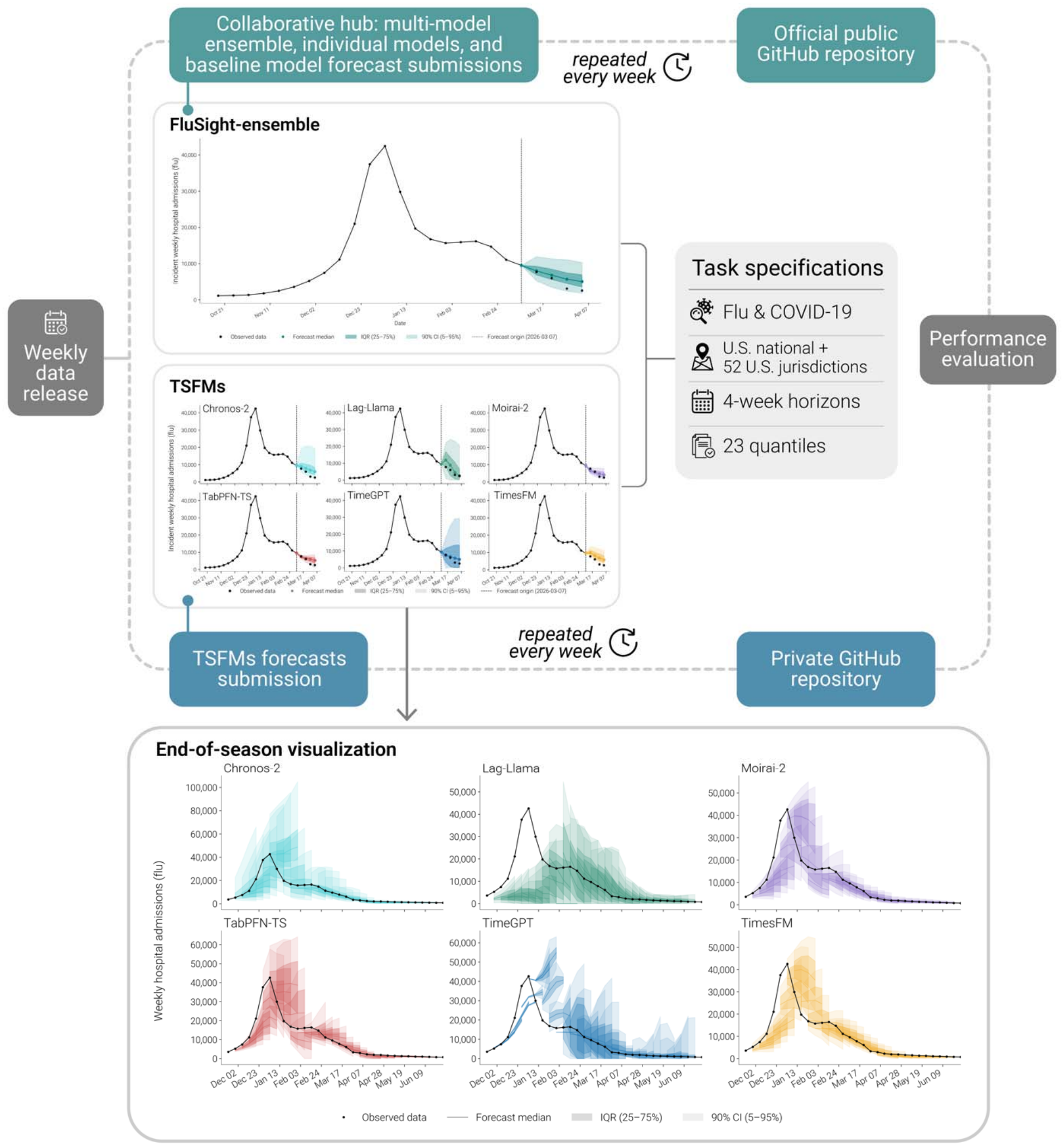
Overview of the epidemiological forecasting task and evaluation framework of time series foundation models (TSFMs). Weekly surveillance data are released by the CDC and ingested by six TSFMs (Chronos-2, Lag-Llama, Moirai-2, TabPFN-TS, TimeGPT, and TimesFM) and by the pool of individual models officially participating in the FluSight and CovidHub hubs. Each week, TSFMs’ probabilistic forecasts are submitted to a private repository. Forecasts from the official individual models participating in the hubs are aggregated into the multi-model ensemble and subsequently released to the official public GitHub repositories. Forecast submissions cover the U.S. national level and 52 U.S. jurisdictions, a 4-week horizon, and 23 quantiles. Forecasts have been submitted weekly over 27 forecasting rounds (November 22, 2025 – May 23, 2026). At the end of the season, forecast performance is evaluated using the weighted interval score (WIS), absolute error (AE), and prediction interval coverage. The pipeline is applied independently to influenza hospital admissions (FluSight) and COVID-19 hospital admissions (CovidHub).

We evaluate six TSFMs representative of the current state of the art in the field: Chronos-2 (Amazon),^38^ Lag-Llama (Mila-Quebec AI Institute),^39^ Moirai-2 (Salesforce),^40^ TabPFN-TS (Prior Labs),^41^ TimeGPT (Nixtla),^42^ and TimesFM (Google).^43^ A detailed description of each model is provided in Sec. 4.1. TSFMs’ performance was evaluated using the weighted interval score (WIS), which jointly captures forecast accuracy, sharpness, and calibration of prediction intervals; absolute error (AE), which considers the deviation of median forecasts from observed values; and prediction interval coverage, which captures the calibration of forecasts as the proportion of observations falling within a specific prediction interval. For each hub, we compared the TSFMs performance to that of contributing models, the ensemble combining individual contributions, and a baseline persistence model. Full methodological details are reported in Section 4.3.

### 2.2 Performance of TSFMs against the FluSight and CovidHub ensemble and individual models

First, we evaluate the performance of the six TSFMs by comparing them with models that submitted forecasts to the FluSight and CovidHub challenges during the 2025/26 season. We focus on the relative log WIS and relative log AE, computed using the pairwise comparison approach proposed in Ref. 26, as well as the 50% and 90% prediction coverage. A relative log WIS/AE score smaller than 1 indicates better performance, whereas a score greater than 1 indicates worse performance relative to the specific hub baseline model. The baseline is a persistence-naive model in both hubs; specifically, a flat forecast equal to the most recently observed value, with prediction intervals derived from historical week-to-week fluctuations (see Sec. 4.2 for further details). Both metrics are computed on log-transformed forecasts and observations.^44^ Coverage values closer to the nominal levels of 0.50 and 0.90 indicate better-calibrated prediction intervals (see Sec. 4.2 and 4.3 for full details on hub specifications and evaluation metrics).

For FluSight, 55 models submitted weekly forecasts of confirmed influenza hospitalizations, including the hub-generated ensemble and baseline models. Following the FluSight inclusion criterion for evaluation,^33^ which requires submission of at least 75% of the forecast instances, 41 models qualified for the analysis, while 14 were excluded. Including the six TSFMs evaluated in this study, the total evaluation set consisted of N = 47 models. For CovidHub, 17 models submitted weekly forecasts of confirmed COVID-19 hospitalizations, including the ensemble and baseline models. Applying the same inclusion criterion, 11 qualified and 6 were excluded, resulting in a total evaluation set of N = 17 models, including the six TSFMs. The full list of eligible and excluded models is reported in the Supplementary Information (Sec. S1).

Fig. 2 reports the performance aggregated across all U.S. locations (national level and 52 jurisdictions) of the six TSFMs, the hub ensemble, and the baseline model, for FluSight (Panel A) and CovidHub (Panel B). We report, in parentheses, the rank of each model relative to the full set of evaluated models for each metric considered. We stress that the hub ensemble was constructed using all participating models that opted in to the ensemble and does not include the six TSFMs evaluated here. The comparison including all individual models is provided in the Supplementary Information (Sec. S2). To contextualize TSFMs’ performance, we classified models into tertiles based on their rank across all evaluated models (see Sec. 4.3). Models were accordingly grouped into top, middle, and bottom tertiles, indicated in the table by blue, grey, and orange cell shading, respectively.

**Figure 2.**
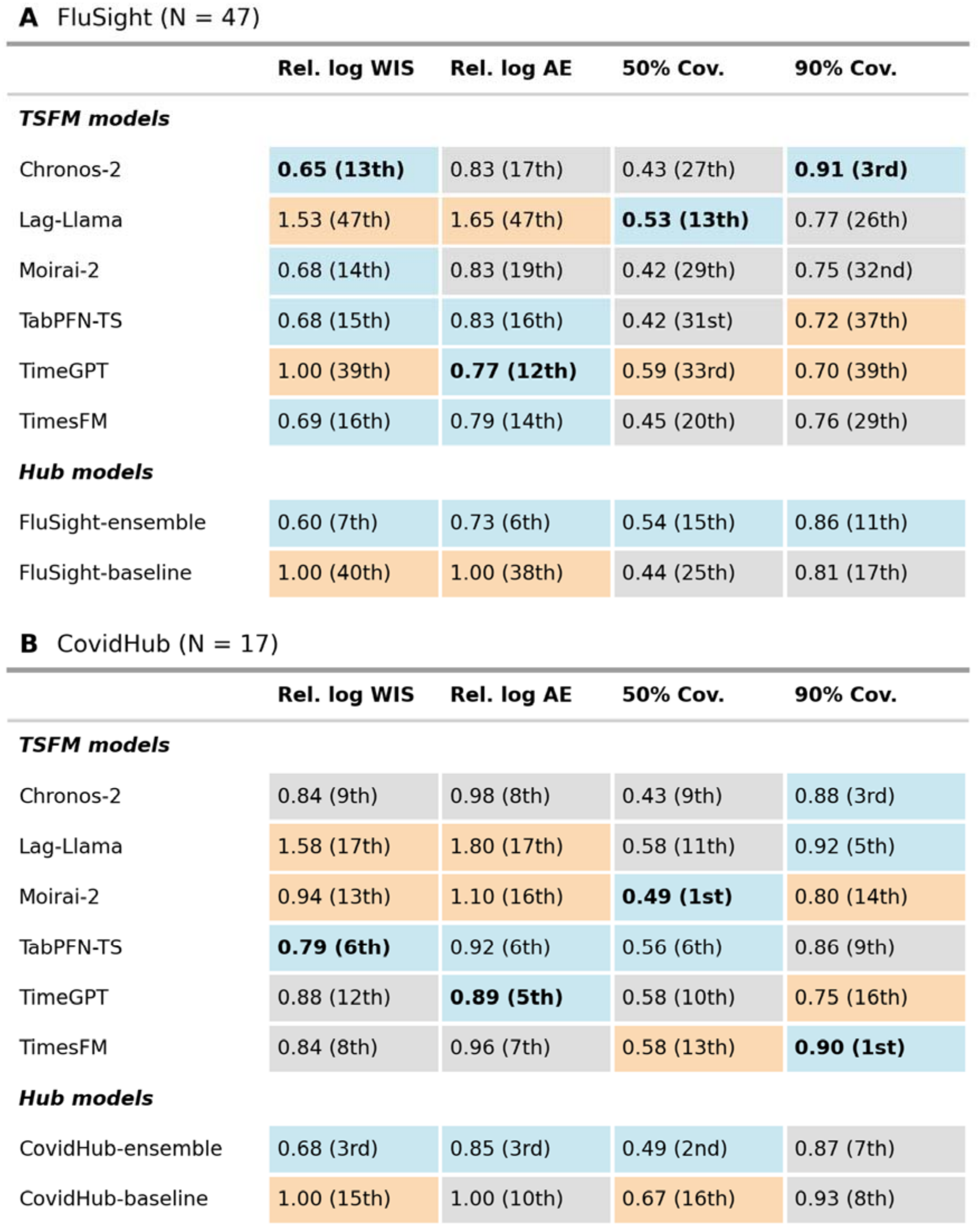
Aggregated forecasting performance of TSFMs and hub models across challenges. Panels show results for (A) FluSight and (B) CovidHub. Performance is reported in terms of relative log WIS (Rel. log WIS), relative log AE (Rel. log AE), and 50% and 90% prediction interval coverage (Cov.). For each metric within eac panel, the best-performing model among the TSFMs is highlighted in bold. For each challenge, N represents the total number of models included in the evaluation, comprising both the models that submitted forecasts to the hu and the six TSFMs evaluated here; rankings in parentheses reflect each model’s position across all N models, with blue, grey, and orange cell shading indicating the top, middle, and bottom tertile, respectively. Note that N reflects the number of eligible models that submit forecasts and may differ from the number of models designated for inclusion in the hub’s official ensemble. TSFMs, time series foundation models; WIS, weighted interval score; AE, absolute error.

Considering FluSight, all TSFMs, except for Lag-Llama, show a relative log WIS and AE smaller than 1, indicating better performance compared to the baseline according to these metrics. Chronos-2 consistently ranked in the top tertile for relative log WIS (13th) and 90% coverage (3rd), while it placed in the middle tertile for relative log AE (17th) and 50% coverage (27th). TimesFM performed in the top tertile for relative log WIS (16th) and relative log AE (14th), and in the middle tertile for 50% and 90% coverage (20th, 29th). TimeGPT ranked in the top tertile for relative log AE (12th), but fell in the bottom tertile for relative log WIS (39th), 50% and 90% coverage (33rd, 39th), indicating inconsistent performance across metrics. Moirai-2 and TabPFN-TS consistently ranked near tertile boundaries, at the top-middle border for relative log WIS and relative log AE, and at the middle-bottom border for coverage. Lag-Llama consistently ranked as the worst-performing model for both relative log WIS and relative log AE (47th), while achieving good 50% coverage (13th). In comparison, the FluSight ensemble ranked consistently in the top tertile across all metrics (7th for relative log WIS, 6th for relative log AE, 15th for coverage at 50%, and 11th for coverage at 90%).

Considering CovidHub, we observe similar heterogeneous performance patterns. TabPFN-TS and Chronos-2 consistently ranked in the top or middle tertile across metrics, never falling into the bottom tertile, while TimesFM fell into the bottom tertile only for 50% coverage. TabPFN-TS ranked in the top tertile for relative log WIS (6th), relative log AE (6th), and 50% coverage (6th), and in the middle tertile for 90% coverage (9th). TimesFM ranked in the middle tertile for relative log WIS (8th) and relative log AE (7th), in the bottom tertile for 50% coverage (13th), and in the top tertile for 90% coverage (1st). Chronos-2 placed in the middle tertile for relative log WIS (9th), relative log AE (8th), and 50% coverage (9th), and in the top tertile for 90% coverage (3rd). TimeGPT ranked in the top tertile for relative log AE (5th), in the middle tertile for relative log WIS (12th) and 50% coverage (10th), and in the bottom tertile for 90% coverage (16th). Lag-Llama ranked in the bottom tertile for both relative log WIS and AE (19th), but in the top and middle tertiles for coverage metrics. Moirai-2 consistently ranked in the bottom tertile across most metrics, showing worse results for COVID-19 than for influenza. The CovidHub ensemble ranked in the top tertile for relative log WIS (3rd), relative log AE (3rd), and coverage at 50% (2nd), and in the middle tertile for coverage at 90% (7th).

### 2.3 TSFMs performance across U.S. jurisdictions

Fig. 3 presents the subnational disaggregated performance analysis, characterizing how each TSFM and the hub ensemble performed across individual locations in terms of relative log WIS and AE. Panels A and C show the distribution of relative log WIS in each location for FluSight and CovidHub, respectively, for the TSFMs, ensemble, and baseline, sorted by ascending median value. The corresponding figure for relative log AE is reported in panels B and D. Each dot represents the relative log WIS (or AE) for a specific U.S. jurisdiction. The empty square marker indicates the value for U.S. national forecasts, and the median across all jurisdictions for each model is highlighted with a diamond-filled marker. The bars on the right of each panel show the number of jurisdictions for which each model ranks first among the models considered here. In the Supplementary Information, we report the relative log WIS and AE values for each jurisdiction (Figs. S1-S2). Unless otherwise specified, relative log WIS and AE values reported in this section are aggregated across all forecast horizons.

**Figure 3.**
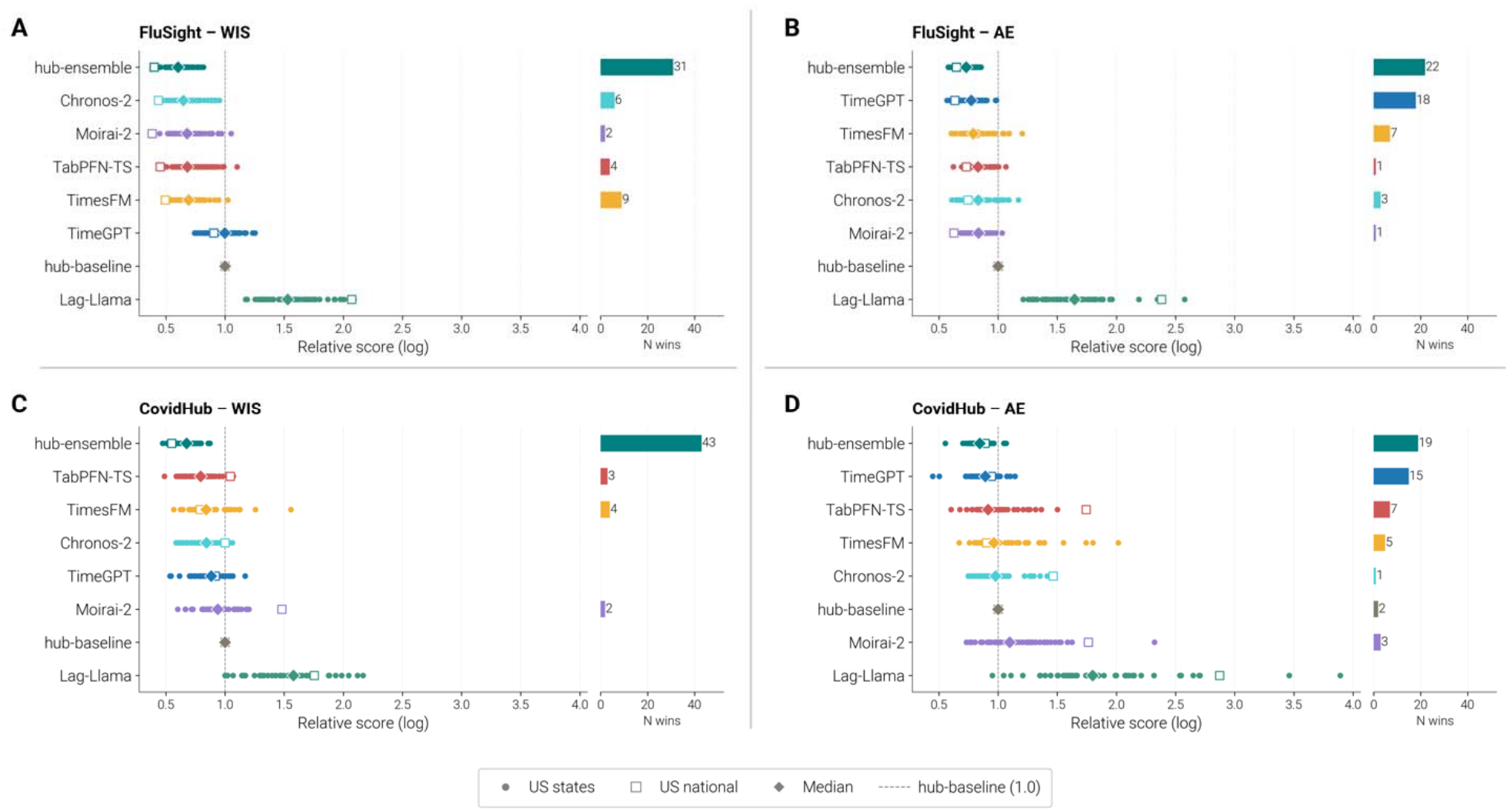
TSFMs performance across locations, 2025/26 season. Relative log WIS and AE of TSFMs and hub models across U.S. jurisdictions and the national level for FluSight (A, B) and CovidHub (C, D). Each dot represents a jurisdiction, the square marker represents the U.S. national, and the diamond indicates the media across all locations. The bars on the right (N wins) represent the number of jurisdictions in which the model ranks first (i.e., achieves the lowest score value). Models are ordered by increasing median. TSFMs, time series foundatio models; WIS, weighted interval score; AE, absolute error.

Focusing on FluSight (Figs. 3A, 3B), models fall into three broad groups. Chronos-2, Moirai-2, TimesFM, and TabPFN-TS consistently outperform the baseline, with median relative log WIS and AE below 1 in most locations. Lag-Llama systematically underperforms, with both metrics above 1 across all locations. TimeGPT exhibits a mixed pattern: it outperforms the baseline in only half of locations for relative log WIS but in all locations for relative log AE, suggesting stronger point forecast accuracy than interval sharpness and calibration. The FluSight ensemble achieves the lowest median relative log WIS and relative log AE across all locations. Among the six TSFMs, the ensemble, and the baseline, the FluSight ensemble ranks first for relative log WIS in the most jurisdictions (31 out of 52), with TimesFM second (9 out of 52) and Chronos-2 third (6 out of 52). For relative log AE, the ensemble leads in 22 locations, followed by TimeGPT (18 locations), and then TimesFM (7 locations).

Considering the CovidHub (Fig. 3C, 3D), we observe similar patterns across models. The ensemble achieves the lowest median relative log WIS and AE. Chronos-2, TimesFM, and TabPFN-TS outperform the baseline on both metrics, with median values below 1, whereas Moirai-2 shows borderline performance, with median values close to 1. TimeGPT again exhibits the same cross-metric inconsistency observed for FluSight, with lower performance in terms of relative log WIS than for AE. Lag-Llama underperforms systematically, falling below the baseline in nearly all locations for both relative log WIS and AE. Among the six TSFMs, the ensemble and the baseline, the ensemble ranks first for relative log WIS in the greatest number of jurisdictions (43 out of 52). For AE, the ensemble ranks first in 19 locations, followed by TimeGPT (15 locations).

### 2.4 Evaluating performance consistency

We explore forecasting performance consistency by evaluating how often each model ranks among the best- and worst-performing models across individual forecasts. Unlike the relative log WIS and AE used in the previous sections, which summarize performance over all forecast through pairwise comparisons, this analysis ranks models directly on their log WIS (or log AE) within each forecast instance, defined by reference date, location, and horizon. For each instance, we identify the models in the top and bottom 30% of the ranking (approximating the top and bottom tertile cutoffs), and compute, for each model, the percentage of its forecast instances in which it fell in the top 30% and in the bottom 30%. We then build a performance distribution map, a scatter plot in which each point represents a model, with coordinates given by the frequencies of top (y-axis) and bottom (x-axis) 30% placements. The median values of coordinates along both axes define four quadrants. Models in the top-left quadrant (Top performers) consistently achieve a low proportion of bottom-ranked forecasts while frequently appearing among the top performers. Models in the bottom-left quadrant (Consistent performers) show low frequency in both extremes, indicating moderate but consistent performance. Model in the top-right quadrant (Inconsistent performers) appear frequently in both the top and bottom rankings, reflecting high variability and inconsistent performance across rounds and locations.

Finally, models in the bottom-right quadrant (Frequent low performers) consistently achieve a low proportion of top-ranked forecasts and frequently rank among the weakest performers.

Fig. 4 shows the performance distribution map for FluSight (first row) and CovidHub (second row) based on log WIS (4A, 4C) and log AE (4B, 4D). Colored circles indicate the TSFMs; grey circles represent other individual models; the square marker represents the baseline model; and the diamond indicates the ensemble.

**Figure 4.**
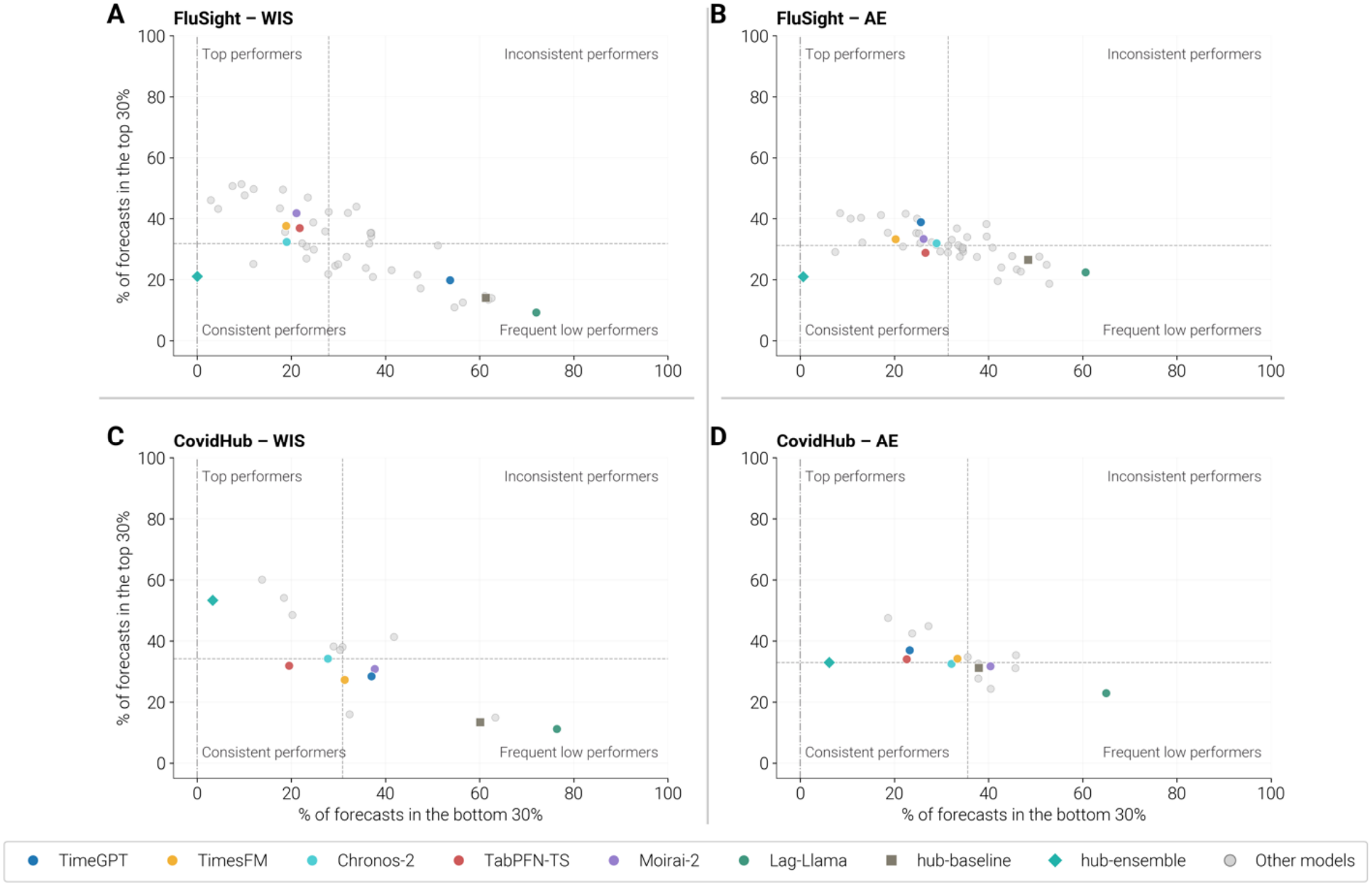
Distribution of top- and bottom-ranked forecasts for FluSight (first row) and CovidHub (second row) during the 2025/26 season. Performance is computed based on log WIS (A, C) and log AE (B, D) rankings. The x-axis shows the percentage of forecasts in which a model ranked in the bottom 30%, while the y-axis shows the percentage of forecasts in which the model ranked in the top 30%. Each colored circle represents a specific TSFM, the diamond represents the ensemble model, the square represents the baseline model, and grey circles represent individual models. Quadrants are defined using the median values of both axes across all models, defining the following categories: Top performers (low bottom-30% frequency, high top-30% frequency), Consistent performers (low frequency in both bottom-30% and top-30%), Inconsistent performers (high frequency in bot bottom-30% and top-30%), and Frequent low performers (high bottom-30% frequency, low top-30% frequency). TSFM, time series foundation model; WIS, weighted interval score; AE, absolute error.

For both diseases and metrics, models cluster along the anti-diagonal, with few models in the top-right corner, indicating that high inconsistency is uncommon. Focusing on FluSight (4A, 4B), TimesFM is in the top-left quadrant (Top performers), ranking in the bottom 30% in approximately 20% of forecasts while appearing in the top 30% in approximately 40% of forecasts for log WIS, and a lower number of forecasts in the top 30% for log AE (nearly 30%). Chronos-2 follows a similar pattern, though with a marginally higher bottom-rank frequency for log AE and a slightly lower top-rank proportion (around 30% for log WIS), positioning it on the boundary between the Top performers and the Consistent performers. For log WIS, Moirai-2 and TabPFN-TS patterns are similar to TimesFM and Chronos-2, falling in the top-left quadrant with high top-rank and low bottom-rank frequencies. For log AE, both models move toward the central region of the plot, indicating more moderate and mixed performance. TimeGPT is placed in the Frequent low performer quadrant for log WIS and in the top-left quadrant as a Top performer for log AE. The baseline model and Lag-Llama are consistently positioned in the Frequent low performer quadrant, with Lag-Llama approaching the bottom-right corner, reflecting a very high proportion of bottom-ranked forecasts. The ensemble is placed in the moderate quadrant for both log WIS and AE, yet stands out for its stability: it never ranks in the bottom 30%, despite not frequently reaching the top 30% as other models. Other individual models are scattered across all quadrants, with a subset reaching the top-left quadrant, indicating that some hub participants achieve consistently strong performance, comparable to or higher than that of the best-performing TSFMs. The CovidHub analysis (4C, 4D) shows consistent patterns. Chronos-2 and TabPFN-TS emerge as the most reliable TSFMs, both occupying top-to-moderate quadrants, while the baseline model and Lag-Llama remain the weakest performers. TimeGPT again confirms its metric-dependent performance: it achieves high top-rank frequency and the second-lowest bottom-rank frequency among TSFMs for log AE, yet shows a low-performer profile for log WIS. For CovidHub, TabPFN-TS records the lowest bottom-rank frequency for log WIS across the six TSFMs and a bottom-rank frequency for log AE comparable to TimeGPT, indicating consistent performance on both metrics. In this case, the ensemble achieves a top-rank proportion of more than 50% for log WIS and approximately 33% for log AE, while again rarely appearing in the bottom rankings, further confirming its consistent and reliable performance.

### 2.5 Evaluating performance across epidemic phases

Fig. 5 presents TSFM performance stratified by epidemic phase for FluSight (left-side panel) and CovidHub (right-side panel), evaluated using both log WIS and AE. For each forecast instance, defined by reference date, location, and horizon, we compute the model’s log WIS (or log AE) relative to that of the baseline. Then, we classify the corresponding week into one of five epidemic phases (large decrease, decrease, stable, increase, large increase) based on the week-to-week difference in hospital admission rates (see Sec. 4.4). Figs. 5A and 5B show the week-by-week classification of U.S. jurisdictions by epidemic phase for FluSight and CovidHub, respectively. The upper panel displays the number of jurisdictions assigned to each phase per week, while the lower panel shows the U.S. national surveillance curve alongside the individual state-level curves in light grey. Performance stratified by epidemic phase is reported in Figs. 5C-F. In the figure, each dot represents the median score ratio for one phase: blue for large decrease, light blue for decrease, grey for stable, orange for increase, and red for large increase. Values below 1 indicate better performance than the baseline, and values above 1 indicate worse performance than the baseline; the x-axis is on a log scale. TSFMs are shown in alphabetical order; for reference, we also report the median performance of the hub ensemble and the average across all individual contributing models.

**Figure 5.**
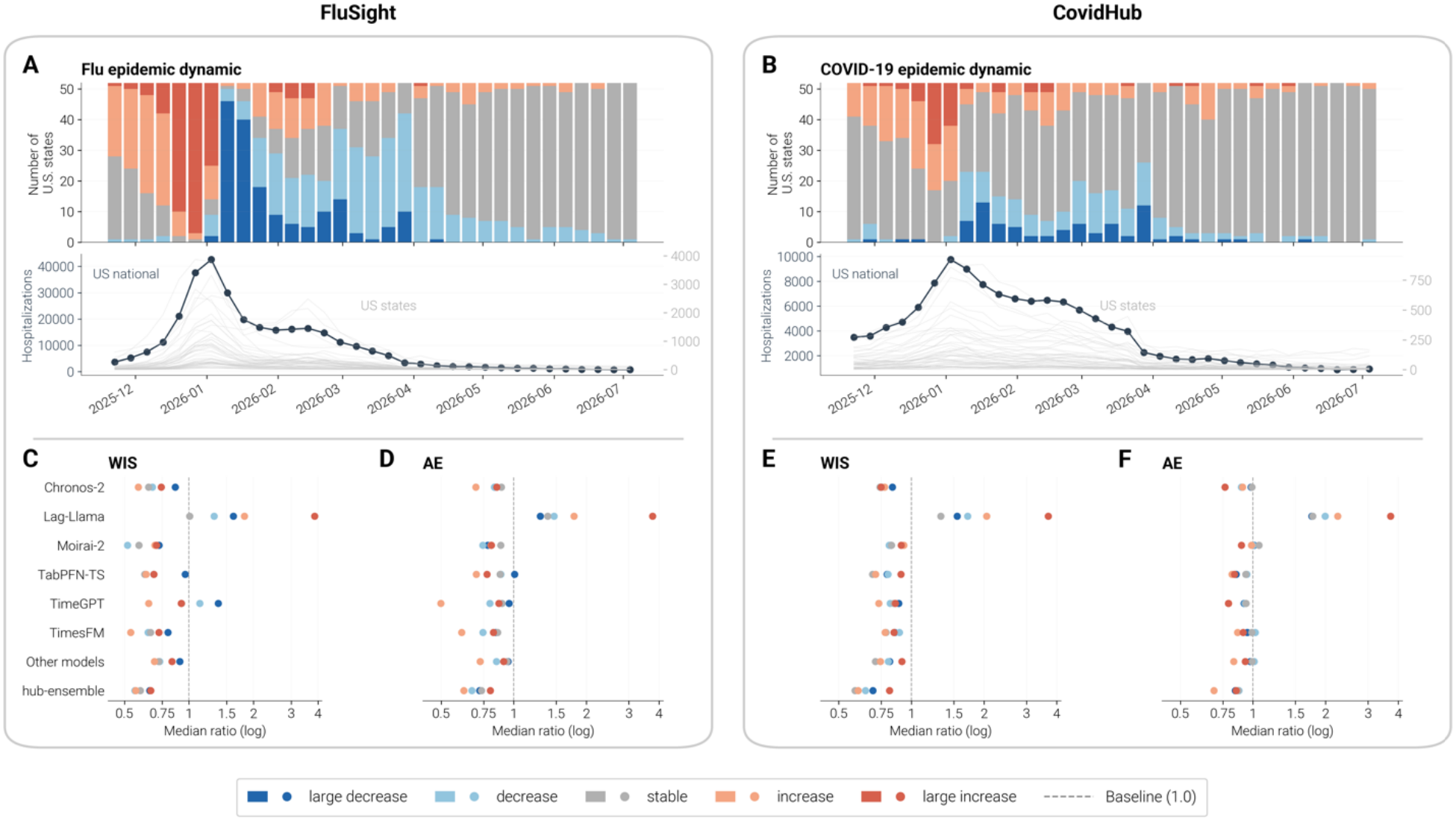
Performance stratified by epidemic phases for FluSight (left panel) and CovidHub (right panel) across the 2025/26 season. Panels A and B show the number of jurisdictions assigned to each phase per week (to row) and the U.S. national surveillance curve alongside the individual jurisdiction-level curves in light grey (bottom row). Panels C-F show performance stratified by epidemic phases based on log WIS (C, E) and log AE (D, F) ratios compared to the baseline. Each dot represents the median score ratio for one epidemic phase: blue for larg decrease, light blue for decrease, grey for stable, orange for increase, and red for large increase. Values to the left of the dashed line (ratio < 1) indicate better performance than the baseline. The x-axis is on a log scale. TSFMs are shown in alphabetical order; for reference, the hub ensemble and the average across all individual contributin models are also reported. TSFMs, time series foundation models; WIS, weighted interval score; AE, absolute error.

Focusing on FluSight (Figs. 5C, 5D), most models perform better during the increase and stable phases, as indicated by the leftward shift of the orange and grey dots relative to the blue ones. This pattern holds across both TSFMs and hub participants, with Lag-Llama showing only a median ratio close to 1 in the stable phase for log WIS, and consistently above 1 across all other phases. TimeGPT shows notable phase-dependent inconsistency, especially for log WIS, performing strongly during the increase and large increase trends but falling below the baseline during decrease and large decrease trends. The ensemble remains the best-performing model across most epidemic phases: in terms of log WIS ratio, it reaches the first position for large increase and large decrease phases, the second position for the stable and decrease trends, where Moirai-2 outperforms it, and again the second position for the increase trend, following TimesFM; in terms of log AE ratio, it reaches the first position for the stable, decrease, and large decrease trends, the second position for large increase, following TabPFN-TS, and the third position for increase, following TimeGPT and TimesFM.

Considering the CovidHub (Figs. 5E, 5F), similar phase-dependent patterns emerge, though the differences are less pronounced, as reflected by the closer clustering of the dots. This may partly reflect the smaller number of weeks classified as non-stable for COVID-19 relative to influenza (see Fig. 5B, top panel). For this reason, phase-specific medians do not diverge as clearly as in the FluSight analysis. The ensemble’s outperformance is also more evident in this case, with its data point consistently positioned to the left of all other models across all epidemic phases, except for the large-increase trend. These results suggest that, when performance is disaggregated by epidemic phase, TSFMs do not outperform the ensemble and behave similarly to the other individual models contributing to the hub.

### 2.6 Retrospective evaluation of the 2024/25 season

To assess the robustness of our findings beyond a single epidemic season, we conducted a retrospective evaluation of the six TSFMs during the 2024/25 season under operational real-time forecasting conditions, which required using versioned data available at the time each forecast was made. Weekly forecasts were produced over 25 forecasting rounds, from November 30, 2024, through May 24, 2025, for both hubs. During this season, 48 models contributed to FluSight and 19 to CovidHub, with 38 and 10 models, respectively, eligible for evaluation. Full details and results are reported in the Supplementary Information (in Sec. S1 and Sec. S3). The overall performance patterns are broadly consistent with those observed for 2025/26. Chronos-2, TabPFN-TS, and TimesFM remain the most competitive TSFMs, never ranking in the bottom tertile for WIS or AE, and consistently outperforming the baseline across most U.S. jurisdictions for influenza and, in terms of WIS, for COVID-19. Chronos-2 performs best for influenza and TabPFN-TS for COVID-19. Lag-Llama ranks last among all evaluated models for both WIS and AE in both hubs. TimeGPT confirms its cross-metric inconsistency, with stronger point-forecast accuracy (AE) than probabilistic performance (WIS), particularly for influenza. Moirai-2 shows moderate performance across different settings. One notable difference from 2025/26 is the phase-dependent pattern: in 2024/25, models tended to perform better in decrease-trend phases than in increase-trend phases, especially for influenza, reversing the pattern observed in 2025/26 and suggesting season-specific differences in the forecast ability of each epidemic phase. Across both seasons and all analyses, the ensemble remains the strongest and most reliable forecasting strategy.

## 3 Discussion

In this work, we systematically evaluated six state-of-the-art TSFMs in an operational setting for short-term epidemic forecasting. The evaluation covered two respiratory disease targets, influenza and COVID-19 hospitalizations, and 53 locations (the U.S. national level and 52 U.S. jurisdictions). TSFMs were benchmarked against multi-model ensembles and individual models, including mechanistic, statistical, machine learning, and deep learning approaches. In addition, we assessed the robustness of our findings through a retrospective performance assessment of the 2024/25 season (Supplementary Information, Sec. S3).

TSFMs represent a potential opportunity for operational epidemic forecasting, offering ready access to state-of-the-art approaches without requiring the specialized expertise needed to build and maintain mechanistic or custom deep learning models. However, how their accuracy compared with that of the disease-specific models explicitly developed for this task remained unclear. Our analyses showed that the performance of the considered TSFMs is highly heterogeneous. Results suggest distinct performance profiles: Chronos-2, TimesFM, and TabPFN-TS emerge as the most competitive and reliable TSFMs across settings; TimeGPT shows metric-dependent performance, achieving strong point forecast accuracy but limited probabilistic calibration; Moirai-2 shows context-dependent and moderate performance, competitive in some settings but insufficiently reliable overall; and Lag-Llama consistently underperforms across disease targets and locations, making it an unreliable forecasting tool in its current form. Performance stratified by epidemic phase further shows that TSFMs share the same phase-dependent strengths and limitations observed in other forecasting approaches. The retrospective analysis of the 2024/25 season is consistent with results observed for 2025/26, supporting the robustness of our findings beyond a single epidemic season, while also highlighting that phase-specific performance can vary from season to season. Importantly, no single TSFM consistently outperforms all individual models and ensembles across all settings. This heterogeneity reflects what is routinely observed among the individual models submitted to forecasting hubs each week, where performance is similarly variable across contributing models; in this sense, TSFMs do not yet represent a paradigm shift but rather a new class of tools that can contribute to the field. This indicates that, at their current stage of maturity, these models cannot be a sole substitute for existing multi-model forecasting efforts. That said, at least some of them show competitive performance and can complement and facilitate current forecasting approaches. As with any forecasting tool, however, one should not treat TSFM outputs as inherently reliable: ongoing evaluation should be a standard part of any workflow that incorporates them. Continued season-to-season assessment will help track the impact and improvements of model updates as they are released, and how year-to-year variability in respiratory illness activity and severity affects performance. Efforts to incorporate the results of real-time evaluations like these into their training and development should be prioritized to improve the operational utility of these tools. This need for evaluation extends beyond recurrent respiratory disease forecasting, as TSFMs are general-purpose models that could also be applied to future health emergencies.

Beyond performance heterogeneity across models, we also observed structural issues in the outputs of some TSFMs, which required post-hoc correction before forecasts could be evaluated. Because several models do not natively produce all 23 required quantiles, interpolation was needed to derive the missing quantile levels. Most TSFMs produced negative values in some instances, which were set to zero. Additionally, a few TSFMs produced quantile forecasts that occasionally violated monotonicity, which we corrected by replacing flagged quantiles via linear interpolation between the nearest valid neighbors. These structural issues indicate that the raw outputs of some TSFMs are not yet directly suited for operational epidemic forecasting without additional post-processing. This is a practical concern: less technical users may not recognize or correct such issues, and could unknowingly deploy these outputs as-is, undermining downstream decisions.

Consistent with the official seasonal hub evaluations,^33^ all scores were computed on the log scale following Bosse et al.^44^. Nonetheless, model rankings and relative scores reported here may differ from those in the published FluSight seasonal evaluation^33^ for the following reasons. First, the pairwise comparison approach scores each model relative to all other models in the evaluation set. Adding the six TSFMs to the pool of hub models therefore changes the relative scores of every model, including the hub models. Second, our evaluation includes Puerto Rico, which is excluded from the official FluSight evaluation. Third, we define each forecast instance by reference date, location, and horizon; differences in how forecasts are grouped may affect the resulting relative scores.

This work comes with limitations. First, we evaluated TSFMs exclusively in a zero-shot setting, providing past data as context and without any task-specific fine-tuning. Indeed, our goal was to characterize out-of-the-box performance. Nonetheless, future work should explore to what extent domain-specific fine-tuning can further improve performance. Relatedly, future work could also explore whether TSFMs can be trained or fine-tuned to improve phase-specific performance, for instance by prioritizing performance during key epidemic phases (e.g., pre-peak). The fact that some of the best-performing TSFMs in our evaluation showed comparatively strong performance during the growth phase suggests this could be a promising avenue for further development. Future work could also explore new forecasting targets beyond weekly hospital admissions, such as epidemic peak timing or growth rate. Also, the TSFMs evaluated here were provided only with NHSN hospital admissions data as context, without incorporating other surveillance indicators, such as emergency department visit data, that are included in some existing modeling approaches and could improve forecast accuracy. Future work should explore whether providing TSFMs with additional indicator data as covariates, where supported, can improve their forecasting performance. It is worth keeping in mind that we focused on six widely used and state-of-the-art TSFMs. However, this is a rapidly expanding field, and a broader set of models will be increasingly available. For this reason, future analyses should expand our performance evaluation to include newer versions and model architectures.^45^ Finally, we acknowledge recent advances in a dedicated line of research aimed at developing domain-specific foundation models for epidemic forecasting.^46–48^ While we deliberately focused on evaluating general-purpose TSFMs, a natural direction for future work is to evaluate these domain-specific foundation models and benchmark them against general-purpose TSFMs, individual hub contributors, and ensembles in an operational forecasting setting.

Overall, our results suggest that current TSFMs are not a single solution for operational short-term epidemic forecasting. Collaborative modeling efforts and multi-model ensembles remain the strongest and most reliable strategy for communicating short-term forecasts and scenario projections to key stakeholders. This work provides early information for the epidemic modeling community on the potential and limitations of frontier TSFMs in operational forecasting settings and highlights the need for rigorous evaluation to make the most of their full potential and effectively integrate them into collective forecasting efforts.

## 4 Methods

Here, we describe the TSFMs selected for evaluation and key implementation details relevant to the epidemic forecasting task, followed by an overview of the collaborative forecasting hubs, the metrics used to evaluate their performance, and the methodologies used to further characterize and stratify these results. Although we do not formally submit forecasts to the hubs, we operate in the same real-time fashion, generating predictions weekly as new data become available. Each TSFM receives as input the historical time series of influenza and COVID-19 hospital admissions starting from the 2020/21 season. A pre-processing step was applied to models that required a complete, gap-free input sequence, filling any missing values via linear interpolation; a post-processing step was applied to models whose quantile outputs occasionally violated monotonicity, correcting them post hoc. An additional post-processing step was applied to models that did not natively produce all 23 required quantile levels; for those models, the missing quantiles were derived via interpolation. Finally, all models except TimesFM produced some negative quantile values, which were clipped to zero. The Supplementary Information (Sec. S4) provides full details on which models were affected by each pre- and post-processing step. All forecasts have been made publicly available at https://github.com/sfiandrino/epi-tsfm-evaluation.

### 4.1 Time series foundation models description

#### Chronos-2

is a time-series foundation model based on an encoder-only Transformer derived from the T5 architecture,^49^ introduced by Amazon Web Services in 2025.^38^ Chronos-2 is trained on a mixture of real-world and synthetic datasets. It incorporates temporal attention to capture long-range dependencies and seasonality and employs a group attention mechanism to share information across similar time series. Chronos-2 directly outputs quantile forecasts over multiple horizons, and this output format is well aligned with the probabilistic forecasting requirements of epidemic modeling. Since Chronos-2 quantile outputs occasionally violate monotonicity, we apply a post hoc correction in which flagged quantiles are replaced by linear interpolation between the nearest valid neighbors. We use the *chronos-forecasting* package (v2.2.1) with the pretrained model amazon/chronos-2.

#### Lag-Llama

is an open-source TSFM for time series forecasting with official model releases in February 2024, based on a decoder-only autoregressive Transformer architecture designed for univariate probabilistic forecasting.^39^ Its key design feature is the use of lagged values as input features, allowing each prediction to incorporate information from selected past observations beyond the most recent context. Predictions are generated via step-by-step autoregressive sampling, and the Transformer representations are passed to a Student’s t-distribution head, from which quantile forecasts are obtained via sampling. The number of samples is a user-defined parameter; we set it to 100 to balance quantile estimate stability and computational feasibility in the operational forecasting context. We use the pretrained lag-llama model, downloaded from the Hugging Face Hub (*time-series-foundation-models/Lag-Llama*).

#### Moirai-2

is a TSFM introduced by Salesforce in 2025, based on a decoder-only autoregressive Transformer architecture.^40^ Moirai-2 employs autoregressive multi-quantile decoding, propagating uncertainty across prediction steps rather than collapsing forecasts to a single value. It outputs quantile forecasts optimized via the quantile (pinball) loss,^40^ providing a theoretically grounded approach to probabilistic forecasting that aligns with the continuous ranked probability score (CRPS). We note that Moirai-2 natively outputs 9 quantile levels (0.1, 0.2, …, 0.9), which is a subset of the 23 quantile levels required by the hub submission format. The remaining quantile levels were obtained by linear interpolation using the 9 available quantiles. Missing values in the input time series are imputed via linear interpolation before model input.^40^ Since Moirai-2 quantile outputs occasionally violate monotonicity, we apply a post-hoc correction in which flagged quantiles are replaced via linear interpolation between the nearest valid neighbors. We use the Moirai2Forecast model from the *uni2ts* package (Salesforce AI Research).

#### TabPFN-TS

is a tabular foundation model and differs from the sequence-based architecture of conventional TSFMs, introduced by Prior Labs in 2024, and officially released in 2025.^41,50^ It applies the regression variant of TabPFN-v2^51^ to a tabular representation of the series built from explicit temporal features, including a running index, calendar-based variables, and automatically inferred seasonal components. Rather than learning temporal structure implicitly from raw sequences, the model relies on engineered features, offering a complementary forecasting approach to the other TSFMs evaluated in this study. TabPFN-TS supports probabilistic forecasting through direct quantile prediction, making it compatible with the submission format required by epidemic collaborative efforts. We note that TabPFN-TS natively outputs 9 quantile levels (0.1, 0.2, …, 0.9), which is a subset of the 23 quantile levels required by the hub submission format. The remaining quantile levels were obtained by linear interpolation using the 9 available quantiles. The input representation is constructed using three temporal feature transformers: a running index, calendar-based features, and automatically inferred seasonal features. Missing values are imputed via linear interpolation before model input, as the architecture requires a complete, gap-free sequence.^50^ We use the *tabpfn-time-series* package (v1.0.7).

#### TimeGPT

is a transformer-based TSFM developed by Nixtla in 2023, based on an encoder-decoder architecture with stacked layers, residual connections, and layer normalization.^42^ TimeGPT is not fully open source; available descriptions indicate that the model was pretrained on 100 billion time-series observations spanning diverse temporal structures, including multiple seasonal patterns, irregular cycles, and varying noise and outlier intensity.^42^ For uncertainty quantification, TimeGPT supports either direct quantile forecasting or prediction intervals via conformal prediction. Here, we extract all 23 quantile levels required by the hub submission format directly via the API’s quantiles parameter, returning the corresponding percentiles of the predictive distribution, which align with the probabilistic forecasting requirements of epidemic modeling. Missing values are imputed via linear interpolation before model input, as the architecture requires a complete, gap-free sequence.^42^ We use the *nixtla* package accessed via the TimeGPT API.

#### TimesFM

is a decoder-only TSFM developed by Google Research for zero-shot forecasting and released in 2024.^43^ It represents the input series as contiguous, non-overlapping patches and processes them through stacked causal self-attention layers. A key design feature is using an output patch length that exceeds the input patch length, thereby reducing the number of autoregressive iterations required for multi-step-ahead prediction. TimesFM also employs patch masking during training, exposing the model to variable context lengths, a useful property in epidemic forecasting where the amount of available historical data can vary across locations and targets. Probabilistic forecasts are generated through a dedicated output layer trained with quantile loss at each required probability level, directly producing quantile forecasts. TimesFM natively outputs 9 quantile levels (0.1, 0.2, …, 0.9), which is a subset of the 23 quantile levels required by the hub submission format. The remaining quantile levels were obtained by linear interpolation using the 9 available quantiles. We use the *timesfm* package installed from the official Google Research repository.

### 4.2 Collaborative hubs description

A collaborative hub is a consortium of independent research teams organized around a shared forecasting task, defined by a common target variable, time period, set of locations, and forecast horizons, in which each team contributes probabilistic predictions generated by its own model.^52^ These forecasts are combined weekly into a multi-model ensemble, as combining outputs from multiple models leverages a long-standing tradition in forecasting and yields a more robust representation of consensus across different approaches than relying on any single model.^27^ Within this framework, the hub plays a central role in coordinating activities, including the collection, archiving, evaluation, and synthesis of forecasts.^26^ Modeling teams are required to submit weekly forecasts to the centralized hub, following a standardized format to ensure consistent aggregation and comparison across models. Typically, teams provide probabilistic forecasts in the form of quantiles.

In this study, we consider the U.S. collaborative hubs for short-term forecasts. FluSight^36^ collects forecasts of weekly laboratory-confirmed influenza hospital admissions across 53 U.S. locations (50 states, Washington DC, Puerto Rico, and the U.S. national level), typically running from November through May. The COVID-19 Forecast Hub (CovidHub)^37^ collects forecasts of weekly confirmed COVID-19 hospital admissions across the same 53 locations from November, operating on a rolling basis from November 2024 onwards. Confirmed weekly hospital admission data are officially released by the NHSN Weekly Hospital Respiratory Dataset.^35^ The target dataset is updated with new data every Wednesday by midday (pending data availability). To ensure real-time forecasting, all forecasts must be submitted weekly by Wednesday end of day. For both hubs, modeling teams provide national- and jurisdiction-level probabilistic forecasts at 23 predefined quantile levels (0.010, 0.025, 0.050, …, 0.950, 0.975, 0.990), covering 0- to 3-weeks ahead of the reference week. This time window includes the four weeks after the most recent hospital admissions data. Teams are not required to submit predictions for all horizons or locations.

For both hubs, the ensemble model is generated by aggregating valid submissions from all models designated for inclusion in the ensemble, using the hubEnsembles package from the Hubverse framework,^53^ and computing it as the quantile-level median across all eligible quantile-based submissions. The baseline model is neutral and serves as a performance benchmark against which both individual models and the ensemble can be evaluated. It provides a simple reference forecast where the median prediction at all future horizons equals the last observed value, and the predictive distribution is derived from the historical distribution of week-to-week changes in incidence.

### 4.3 Forecast evaluation

#### Scoring scale

Following Bosse et al.^44^, all scores are computed on the log-transformed scale. Before scoring, both forecast quantiles and observed values are transformed as *log(y + 1)*, where the offset avoids undefined values at zero. Scoring on the log scale approximates a measure of relative rather than absolute error. This stabilizes score variance across locations and prevents high-burden jurisdictions from dominating aggregate comparisons. Accordingly, WIS and AE (Eqs. 1-3) and the relative WIS and relative AE derived from them (Eqs. 5-7) are computed on log-transformed quantities, and expressed in the text as log WIS, log AE, relative log WIS and relative log AE.

#### Weighted Interval Score (WIS)

The WIS is a proper scoring rule that evaluates both the accuracy and calibration of probabilistic forecasts. It summarizes how well a forecast distribution captures observed outcomes by combining two properties: sharpness, i.e., the narrowness of the prediction intervals, and calibration, i.e., the agreement between predicted and realized probabilities. Well-calibrated and sharp forecasts achieve lower WIS values. The WIS is a standard tool for comparing probabilistic forecasts in the context of infectious disease ensemble forecasting.^34^

Formally, for a given forecast distribution *F* with predictive median *m*, and *K* central (1−*α*_*k*_)×100% prediction intervals defined by lower and upper bounds *l*_*k*_ and *u*_*k*_, the interval score (IS) at level *α*_*k*_ is defined as:

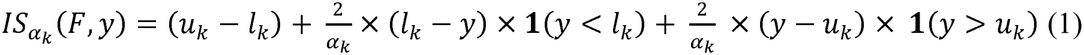

where *y* denotes the observed value and 1{·} is an indicator function.

The overall WIS is then obtained as a weighted average of the absolute error of the predictive median and the interval scores across all considered levels:

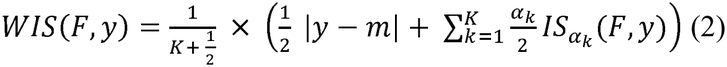

Following standard practice,^34^ we used *K* = 11 central prediction intervals corresponding to *α*_*k*_ = 0.02, 0.05, 0.10, 0.20, 0.30, 0.40, 0.50, 0.60, 0.70, 0.80, and 0.90.

#### Absolute Error (AE)

The AE evaluates point forecasts. For each forecast instance *i*, let *ŷ*_*i*_ denote the predicted median value and *y*_*i*_ the corresponding observed value. The AE of the median measures the magnitude of the absolute deviation between forecast and observation as:

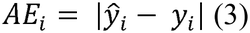

#### Prediction coverage

Prediction interval coverage is a measure of model calibration and represents the proportion of observations that fall within a given prediction interval. For instance, a perfectly calibrated model will have exactly 90% of observations within the 90% prediction intervals. Formally, for a prediction interval of level *α* and a set of *N* observations *y*_*i*_, coverage is defined as:

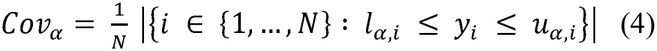

#### Relative performance

We compute the relative performance for WIS and AE using a pairwise comparison approach in which the relative skill of a model *m* with respect to the set of all other models {1, …, M}, including the baseline model, is considered.^26^ For each pair of models *m* and *m*^′^, we compute the pairwise relative WIS and relative AE skill as:

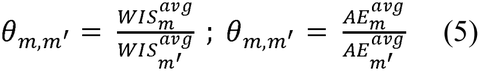

where 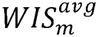 and 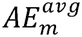 (resp. 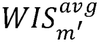 and 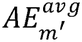) are the average of WIS or AE of the model (resp. ) computed using all available forecasts where both model and model participated. Then, for each model, we compute the geometric mean across the different models:

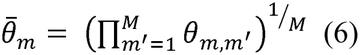

Finally, to compare model performance with respect to the baseline model, we divide 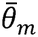 by the result obtained for the baseline, named 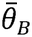:

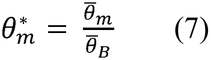

The obtained value 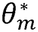 represents the relative WIS or relative AE of model rescaled to the baseline model, such that the baseline has a value of 1. It follows that 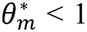 indicates that the model performs better than the baseline, and 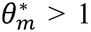 indicates that the model performs worse than the baseline. All scores are computed on the log-transformed scale^44^ as described above.

#### Tertile classification

To contextualize model performance beyond raw relative scores, we additionally classified models into tertiles based on their rank across all *M* models evaluated within each challenge (for both relative log WIS and relative log AE). Models were ranked from best to worst-performing (i.e., from lowest to highest), and divided into three groups of approximately equal size using the 33.3rd and 66.7th percentiles of the rank distribution as cutoffs. The top tertile includes the best-ranked third of models, the middle tertile the next third, and the bottom tertile the remaining, worst-ranked third. When *M* is not evenly divisible by three, the remaining model(s) were sequentially assigned to the higher tertiles (top, then middle), so that the bottom tertile was never larger than the other two.

### 4.4 Disease trend classification

To characterize model performance across different epidemic phases, we developed a method to classify trends in influenza and COVID-19 activity based on historical data. We adapted the methodology originally developed by the CDC for influenza activity,^54^ applying the same approach to both diseases using hospital admission data from NHSN.^35^ We derived weekly rates per 100,000 population and computed the week-to-week difference in these rates for each season, considering all available historical data from the 2022/23 season onward, to exclude COVID-19 seasons in which activity patterns may have been distorted by pandemic period dynamics (2019/20, 2020/21, 2021/22). From the resulting distribution of rate differences, we established five categories of change: large decrease, decrease, stable, increase, and large increase. We derived a single set of thresholds using national-level (U.S.) rate differences as the reference distribution, based on the 5th, 25th, 75th, and 95th percentiles, which we used as cutoffs to define the trend categories. Specifically, differences greater than the 25th percentile and lower than the 75th percentile were classified as stable; differences between the 75th (inclusive) and 95th percentiles were classified as an increase; differences between the 5th and 25th (inclusive) percentiles were classified as a decrease; and differences at or below the 5th percentile or at or above the 95th percentile were classified as a large decrease or large increase, respectively. For each jurisdiction, weeks with an absolute change of <10 hospital admissions were classified as stable, to limit the impact of reporting delays and random fluctuations.

Based on these criteria, for FluSight, stable weeks were defined as a change in weekly rate ≥ −0.09 to < 0.04 influenza-associated hospitalizations/100,000, or as weeks with an absolute difference of <10 hospital admissions. Increases were defined as a positive change in weekly rate ≥ 0.04 to < 1.59/100,000, and large increases as a positive change ≥ 1.59/100,000. Decreases were defined as a negative change in weekly rate > −1.39 to < −0.09/100,000, and large decreases as a negative change ≤ −1.39/100,000. For CovidHub, stable weeks were defined as a change in weekly rate ≥ −0.30 to < 0.12 COVID-19-associated hospitalizations/100,000, or as weeks with an absolute difference of <10 hospital admissions. Increases were defined as a positive change in weekly rate ≥ 0.12 to < 0.86/100,000, and large increases as a positive change ≥ 0.86/100,000. Decreases were defined as a negative change in weekly rate > −0.77 to < −0.30/100,000, and large decreases as a negative change ≤ −0.77/100,000.

In the Supplementary Information (Sec. S5), we present a more detailed visualization of the resulting trend classification applied to the U.S. national curve for both diseases. We also conducted a sensitivity analysis to evaluate our threshold-derivation approach. For influenza, we compared the obtained thresholds and trend classifications against those officially reported by the CDC in ref. 54, which were instead derived from an independent surveillance system (FluSurv-NET). Despite this difference in derivation, agreement between our influenza trend classifications and those obtained using the original CDC thresholds for the 2024/25 and 2025/26 seasons was high, with a percentage of agreement of 89%. We also quantified the agreement using Cohen’s Kappa, a statistic that measures inter-rater agreement while accounting for the level of agreement expected by chance alone. Kappa values above 0.80 are conventionally considered near-perfect agreement. The resulting Kappa of 0.83 supports the validity of our adapted approach for classifying disease trends.

### 4.5 Use of AI tools

We used Claude (Anthropic) to assist with manuscript preparation in the following ways. We used it as a coding assistant for the scripts used to plot the figures. The authors reviewed the plotting scripts and ran them on the study data. No figure or image was generated by AI. Claude was also used for copy editing of author-written text, limited to wording and grammar. No AI tool generated scientific content or produced the analyses, results, or conclusions. We used the time series foundation models subject to the research to produce weekly forecasts on publicly available data. The authors take full responsibility for this article’s content.

## Data Availability

The data to reproduce all the analyses are available at https://github.com/cdcepi/FluSight-forecast-hub/ and https://github.com/sfiandrino/epi-tsfm-evaluation.

## Code Availability

All forecasts are available at https://github.com/sfiandrino/epi-tsfm-evaluation.

## Acknowledgements

The authors gratefully acknowledge the FluSight Influenza Forecast Hub and the COVID-19 Forecast Hub.

## Author Contributions Statement

SF, NG, and AV conceived and designed the study. SF, AU, SV, and NG implemented the models and produced the forecasts. SF performed the analyses. AP, AU, and SV contributed to the methodology and analyses. RKB and MB contributed to the study design and interpretation of results. SF wrote the first draft of the manuscript. NG and AV supervised the project. All authors critically revised the manuscript and approved the final version.

