## Supplementary Information for "Are time series foundation models the new frontier for operational epidemic forecasting?"

### S1 Summary of individual models contributing to short-term collaborative forecasting hubs during the 2024/25 and 2025/26 seasons

The FluSight and CovidHub forecasting hubs are short-term forecasting challenges, in which modeling teams submit weekly forecasts of confirmed influenza and COVID-19 hospitalizations. During the 2024/25 season, 48 models contributed to FluSight and 19 to CovidHub; during 2025/26, 55 and 17, respectively (counts include the hub ensemble and baseline models). Only models that submitted at least 75% of forecast instances were eligible for evaluation. After applying this criterion, 38 FluSight and 10 CovidHub models were evaluated for 2024/25 (10 and 9 excluded), and 41 FluSight and 11 CovidHub models for 2025/26 (14 and 6 excluded). Tables S1 and S2 list the individual contributing models for each season, the model eligibility for end-of-the-season evaluation analyses, and the type of models, classified as statistical, mechanistic, ensemble, or models with deep learning or machine learning components. Among the excluded models, one model submitted forecasts for only a single jurisdiction in FluSight 2024/25, FluSight 2025/26, and CovidHub 2025/26, and therefore did not meet the 75% criterion.

| **Model Name** | **2024/25** | **2025/26** | **Eligibility** | **Model type** |
| --- | --- | --- | --- | --- |
| CADPH-FluCAT_Ensemble | ✓ | ✓ | × | ENS, STAT, DL/ML |
| CEPH-Rtrend_fluH | ✓ | ✓ | ✓ | MECH |
| CFA_Pyrenew-Pyrenew_H_Flu | ✓ | ✓ | × | MECH |
| CFA_Pyrenew-Pyrenew_HE_Flu | ✓ | ✓ | × | MECH |
| CMU-TimeSeries | ✓ | ✓ | ✓ | ENS, STAT |
| CMU-climate_baseline | ✓ | × | ✓ | ENS, STAT |
| CU-ARNB_Net | × | ✓ | ✓ | DL/ML |
| CU-ensemble | ✓ | ✓ | ✓ | ENS, MECH, DL/ML, STAT |
| Cornell_JHU-hierarchSIR | × | ✓ | ✓ | MECH, STAT |
| DMAPRIME-DLM | × | ✓ | × | STAT |
| DMAPRIME-QR | × | ✓ | × | STAT |
| Epistorm-Ensemble_Flu | × | ✓ | × | ENS, STAT |
| FluSight-baseline | ✓ | ✓ | ✓ | STAT |
| FluSight-ensemble | ✓ | ✓ | ✓ | ENS, STAT |
| Gatech-ensemble_point | ✓ | × | ✓ | ENS, DL/ML, STAT |
| Gatech-ensemble_prob | ✓ | ✓ | ✓ | ENS, STAT |
| Gatech-ensemble_stat | × | ✓ | ✓ | ENS, STAT, DL/ML |
| Google_SAI-FluBoostQR | ✓ | × | × | DL/ML |
| Google_SAI-FluEns | × | ✓ | ✓ | ENS, MECH, DL/ML, STAT |
| ISU_NiemiLab-GPE | ✓ | × | ✓ | STAT |
| JHUAPL-DMD | ✓ | × | ✓ | STAT |
| JHUAPL-Morris | ✓ | × | × | DL/ML |
| JHU_CSSE-CSSE_Ensemble | ✓ | ✓ | ✓ (2024/25), × (2025/26) | ENS, DL/ML, STAT |
| LUcompUncertLab-chimera | ✓ | ✓ | ✓ (2024/25), × (2025/26) | MECH |
| LUcompUncertLab-puca | × | ✓ | × | STAT |
| LosAlamos-DoSiDo | × | ✓ | ✓ | STAT |
| LosAlamos-ThinMint | × | ✓ | ✓ | DL/ML |
| LosAlamos_NAU-CModel_Flu | ✓ | ✓ | ✓ | MECH |
| MDPredict-SIRS | ✓ | ✓ | × | MECH |
| MIGHTE-Joint | ✓ | ✓ | ✓ | ENS, DL/ML, STAT |
| MIGHTE-Nsemble | ✓ | ✓ | ✓ | ENS, DL/ML, STAT |
| MOBS-EpyStrain_Flu | × | ✓ | ✓ | MECH |
| MOBS-GLEAM_FLUH | ✓ | × | ✓ | MECH |
| MOBS-GLEAM_RL_FLUH | × | ✓ | ✓ | DL/ML, MECH |
| Metaculus-cp | ✓ | × | × | ENS, STAT |
| NAU-FourCAT | × | ✓ | ✓ | DL/ML |
| NAU-epymorph | × | ✓ | ✓ | MECH |
| NAU-vulPES | × | ✓ | ✓ | ENS, STAT |
| NEU_ISI-AdaptiveEnsemble | ✓ | ✓ | ✓ | ENS, STAT |
| NEU_ISI-FluBcast | ✓ | ✓ | ✓ | MECH |
| NIH-Flu_ARIMA | ✓ | ✓ | ✓ | MECH, STAT |
| NU-PGF_FLUH | × | ✓ | × | MECH, STAT |
| OHT_JHU-nbxd | ✓ | ✓ | ✓ | ENS, DL/ML |
| PSI-PROF | ✓ | ✓ | ✓ | MECH, STAT |
| PSI-PROF_MOA | × | ✓ | ✓ | MECH, STAT |
| PSI-PROF_beta | ✓ | × | ✓ | MECH, STAT |
| SigSci-TSENS | ✓ | ✓ | ✓ | ENS, STAT |
| Stevens-ILIForecast | ✓ | × | ✓ | ENS, STAT |
| UGA_CEID-Walk | ✓ | ✓ | ✓ | STAT |
| UGA_CEID-auto_AVG_LB | × | ✓ | ✓ | STAT |
| UGA_flucast-Copycat | ✓ | ✓ | ✓ | STAT |
| UGA_flucast-INFLAenza | ✓ | ✓ | ✓ | DL/ML, STAT |
| UGA_flucast-Scenariocast | ✓ | ✓ | ✓ | STAT |
| UGuelph-CompositeCurve | ✓ | ✓ | × | STAT |
| UGuelphensemble-GRYPHON | ✓ | ✓ | × | ENS |
| UI_CompEpi-EpiGen | ✓ | ✓ | ✓ | ENS, DL/ML, STAT |
| UM-DeepOutbreak | ✓ | ✓ | ✓ | DL/ML |
| UMass-AR2 | ✓ | ✓ | ✓ | STAT |
| UMass-flusion | ✓ | ✓ | ✓ | ENS, DL/ML, STAT |
| UMass-trends_ensemble | ✓ | ✓ | ✓ | ENS, STAT |
| UNC_IDD-InfluPaint | ✓ | ✓ | ✓ | DL/ML |
| UVAFluX-CESGCN | ✓ | ✓ | ✓ (2024/25), × (2025/26) | DL/ML |
| UVAFluX-Ensemble | ✓ | ✓ | ✓ | ENS, DL/ML, STAT |
| UVAFluX-FS_OptimWISE | × | ✓ | ✓ | ENS, STAT |
| UVAFluX-OptimWISE | ✓ | × | × | ENS, STAT |
| VTSanghani-PRIME | ✓ | ✓ | ✓ | DL/ML |
| fjordhest-ensemble | ✓ | × | ✓ | ENS, MECH, STAT |

**Table S1. Summary of participating models in the 2024/25 and 2025/26 FluSight challenge.** Model type classification: ENS (ensemble), STAT (statistical), MECH (mechanistic), DL/ML (deep learning or machine learning).

| **Model Name** | **2024/25** | **2025/26** | **Eligibility** | **Model type** |
| --- | --- | --- | --- | --- |
| CADPH-CovidCAT_Ensemble | × | ✓ | × | ENS, STAT, DL/ML |
| CEPH-Rtrend_covid | ✓ | ✓ | ✓ | MECH |
| CFA-EpiAutoGP | × | ✓ | ✓ | ENS, STAT |
| CFA_Pyrenew-PyrenewHEW_COVID | ✓ | ✓ | × | MECH |
| CFA_Pyrenew-Pyrenew_HE_COVID | ✓ | ✓ | × | MECH |
| CFA_Pyrenew-Pyrenew_HW_COVID | ✓ | ✓ | × | MECH |
| CFA_Pyrenew-Pyrenew_H_COVID | ✓ | ✓ | × | MECH |
| CMU-TimeSeries | ✓ | ✓ | ✓ | ENS, STAT |
| CMU-climate_baseline | ✓ | × | ✓ | ENS, STAT |
| CovidHub-baseline | ✓ | ✓ | ✓ | STAT |
| CovidHub-ensemble | ✓ | ✓ | ✓ | ENS, STAT |
| Google_SAI-Ensemble | ✓ | ✓ | × (2024/25), ✓ (2025/26) | ENS, DL/ML |
| JHU_CSSE-CSSE_Ensemble | ✓ | × | × | ENS, DL/ML, STAT |
| MOBS-GLEAM_COVID | ✓ | × | ✓ | MECH |
| Metaculus-cp | ✓ | × | × | ENS, STAT |
| NEU_ISI-AdaptiveEnsemble | ✓ | ✓ | × | ENS, STAT |
| OHT_JHU-nbxd | ✓ | ✓ | ✓ | ENS, DL/ML |
| UGA_flucast-INFLAenza | ✓ | ✓ | × (2024/25), ✓ (2025/26) | STAT |
| UM-DeepOutbreak | ✓ | ✓ | ✓ | DL/ML |
| UMass-ar6_pooled | ✓ | ✓ | ✓ | STAT |
| UMass-gbqr | ✓ | ✓ | ✓ | STAT |

**Table S2. Summary of participating models in the 2024/25 and 2025/26 CovidHub challenge.** Model type classification: ENS (ensemble), STAT (statistical), MECH (mechanistic), DL/ML (deep learning or machine learning).

### S2 TSFMs performance against the FluSight and CovidHub ensembles and individual models

Tables S3 and S4 report the aggregated performance across the U.S. locations (U.S. national and 52 jurisdictions) of all the individual models contributing to the collaborative hubs, the hub ensemble, the baseline model, and the evaluated TSFMs for FluSight and the CovidHub, respectively. We report relative log WIS (Rel. log WIS), relative log AE (Rel. log AE), 50% and 90% prediction interval coverage. Models are ranked based on the relative log WIS (Rel. log WIS). As in the main text, all scores are computed on log-transformed forecasts and observations, log(y + 1) (see Methods, Sec. 4.3).

| **Model** | **Rel. log WIS** | **Rel. log AE** | **50% Cov.** | **90% Cov.** |
| --- | --- | --- | --- | --- |
| Google_SAI-FluEns | 0.55 (1st) | 0.67 (1st) | 0.52 (8th) | 0.89 (5th) |
| OHT_JHU-nbxd | 0.56 (2nd) | 0.67 (2nd) | 0.43 (28th) | 0.78 (23rd) |
| CMU-TimeSeries | 0.57 (3rd) | 0.71 (5th) | 0.48 (6th) | 0.88 (7th) |
| UGA_flucast-INFLAenza | 0.58 (4th) | 0.68 (3rd) | 0.50 (1st) | 0.85 (13th) |
| UMass-flusion | 0.58 (5th) | 0.69 (4th) | 0.35 (42nd) | 0.74 (33rd) |
| NAU-vulPES | 0.60 (6th) | 0.75 (9th) | 0.47 (14th) | 0.88 (8th) |
| **FluSight-ensemble** | **0.60 (7th)** | **0.73 (6th)** | **0.54 (15th)** | **0.86 (11th)** |
| UGA_flucast-Scenariocast | 0.60 (8th) | 0.78 (13th) | 0.46 (16th) | 0.85 (12th) |
| UVAFluX-FS_OptimWISE | 0.61 (9th) | 0.74 (8th) | 0.44 (22nd) | 0.82 (16th) |
| MIGHTE-Nsemble | 0.63 (10th) | 0.76 (11th) | 0.52 (9th) | 0.89 (4th) |
| NEU_ISI-AdaptiveEnsemble | 0.64 (11th) | 0.74 (7th) | 0.36 (41st) | 0.68 (40th) |
| MIGHTE-Joint | 0.64 (12th) | 0.76 (10th) | 0.62 (40th) | 0.93 (9th) |
| **Chronos-2** | **0.65 (13th)** | **0.83 (17th)** | **0.43 (27th)** | **0.91 (3rd)** |
| **Moirai-2** | **0.68 (14th)** | **0.83 (19th)** | **0.42 (29th)** | **0.75 (32nd)** |
| **TabPFN-TS** | **0.68 (15th)** | **0.83 (16th)** | **0.42 (31st)** | **0.72 (37th)** |
| **TimesFM** | **0.69 (16th)** | **0.79 (14th)** | **0.45 (20th)** | **0.76 (29th)** |
| NAU-FourCAT | 0.71 (17th) | 0.87 (24th) | 0.46 (17th) | 0.90 (1st) |
| NAU-epymorph | 0.71 (18th) | 0.83 (18th) | 0.33 (43rd) | 0.67 (41st) |
| CU-ARNB_Net | 0.72 (19th) | 0.87 (23rd) | 0.54 (18th) | 0.90 (2nd) |
| PSI-PROF | 0.73 (20th) | 0.87 (25th) | 0.56 (23rd) | 0.91 (6th) |
| UGA_flucast-Copycat | 0.73 (21st) | 0.88 (27th) | 0.38 (38th) | 0.75 (31st) |
| UGA_CEID-auto_AVG_LB | 0.74 (22nd) | 0.85 (21st) | 0.42 (30th) | 0.76 (28th) |
| PSI-PROF_MOA | 0.74 (23rd) | 0.81 (15th) | 0.47 (12th) | 0.75 (30th) |
| Gatech-ensemble_prob | 0.76 (24th) | 0.86 (22nd) | 0.51 (3rd) | 0.79 (21st) |
| MOBS-GLEAM_RL_FLUH | 0.77 (25th) | 0.85 (20th) | 0.24 (45th) | 0.53 (45th) |
| MOBS-EpyStrain_Flu | 0.78 (26th) | 0.90 (30th) | 0.38 (39th) | 0.71 (38th) |
| Cornell_JHU-hierarchSIR | 0.80 (27th) | 1.05 (41st) | 0.39 (37th) | 0.76 (27th) |
| CEPH-Rtrend_fluH | 0.80 (28th) | 1.02 (39th) | 0.42 (32nd) | 0.82 (14th) |
| UVAFluX-Ensemble | 0.81 (29th) | 0.93 (32nd) | 0.45 (21st) | 0.73 (35th) |
| NEU_ISI-FluBcast | 0.82 (30th) | 0.88 (28th) | 0.23 (46th) | 0.48 (46th) |
| Gatech-ensemble_stat | 0.82 (31st) | 0.88 (26th) | 0.52 (7th) | 0.78 (25th) |
| NIH-Flu_ARIMA | 0.85 (32nd) | 0.95 (33rd) | 0.26 (44th) | 0.61 (44th) |
| UNC_IDD-InfluPaint | 0.86 (33rd) | 1.03 (40th) | 0.44 (24th) | 0.78 (24th) |
| CU-ensemble | 0.87 (34th) | 0.99 (36th) | 0.51 (4th) | 0.80 (18th) |
| UMass-AR2 | 0.87 (35th) | 0.99 (37th) | 0.47 (11th) | 0.78 (22nd) |
| VTSanghani-PRIME | 0.90 (36th) | 0.97 (34th) | 0.55 (19th) | 0.73 (36th) |
| UI_CompEpi-EpiGen | 0.91 (37th) | 0.91 (31st) | 0.41 (34th) | 0.65 (43rd) |
| UMass-trends_ensemble | 0.98 (38th) | 0.97 (35th) | 0.41 (35th) | 0.65 (42nd) |
| **TimeGPT** | **1.00 (39th)** | **0.77 (12th)** | **0.59 (33rd)** | **0.70 (39th)** |
| **FluSight-baseline** | **1.00 (40th)** | **1.00 (38th)** | **0.44 (25th)** | **0.81 (17th)** |
| SigSci-TSENS | 1.07 (41st) | 1.28 (45th) | 0.60 (36th) | 0.80 (19th) |
| UM-DeepOutbreak | 1.07 (42nd) | 0.89 (29th) | 0.57 (26th) | 0.82 (15th) |
| LosAlamos-DoSiDo | 1.10 (43rd) | 1.23 (44th) | 0.51 (2nd) | 0.86 (10th) |
| LosAlamos-ThinMint | 1.10 (44th) | 1.41 (46th) | 0.47 (10th) | 0.80 (20th) |
| LosAlamos_NAU-CModel_Flu | 1.12 (45th) | 1.06 (42nd) | 0.19 (47th) | 0.39 (47th) |
| UGA_CEID-Walk | 1.19 (46th) | 1.16 (43rd) | 0.51 (5th) | 0.73 (34th) |
| **Lag-Llama** | **1.53 (47th)** | **1.65 (47th)** | **0.53 (13th)** | **0.77 (26th)** |

**Table S3. Aggregated forecasting performance of all FluSight models and TSFMs (N = 47), season 2025/26.** Columns report relative log WIS (Rel. log WIS), relative log AE (Rel. log AE), and 50% and 90% prediction interval coverage. Rankings in parentheses refer to all models in the challenge; for coverage, rank 1 corresponds to the model closest to the nominal level (0.50 and 0.90, respectively). In bold are the TSFM models, the ensemble, and the baseline. Models are sorted by Rel. log WIS. TSFMs, time series foundation models; WIS, weighted interval score; AE, absolute error.

| **Model** | **Rel. log WIS** | **Rel. log AE** | **50% Cov.** | **90% Cov.** |
| --- | --- | --- | --- | --- |
| Google_SAI-Ensemble | 0.64 (1st) | 0.79 (1st) | 0.42 (12th) | 0.85 (10th) |
| CMU-TimeSeries | 0.67 (2nd) | 0.85 (4th) | 0.47 (4th) | 0.88 (6th) |
| **CovidHub-ensemble** | **0.68 (3rd)** | **0.85 (3rd)** | **0.49 (2nd)** | **0.87 (7th)** |
| UGA_flucast-INFLAenza | 0.69 (4th) | 0.83 (2nd) | 0.44 (7th) | 0.82 (13th) |
| CEPH-Rtrend_covid | 0.79 (5th) | 0.99 (9th) | 0.48 (3rd) | 0.83 (12th) |
| **TabPFN-TS** | **0.79 (6th)** | **0.92 (6th)** | **0.56 (6th)** | **0.86 (9th)** |
| CFA-EpiAutoGP | 0.83 (7th) | 1.00 (11th) | 0.43 (8th) | 0.84 (11th) |
| **TimesFM** | **0.84 (8th)** | **0.96 (7th)** | **0.58 (13th)** | **0.90 (1st)** |
| **Chronos-2** | **0.84 (9th)** | **0.98 (8th)** | **0.43 (9th)** | **0.88 (3rd)** |
| UMass-gbqr | 0.84 (10th) | 1.04 (12th) | 0.37 (14th) | 0.76 (15th) |
| UMass-ar6_pooled | 0.86 (11th) | 1.07 (13th) | 0.54 (5th) | 0.91 (2nd) |
| **TimeGPT** | **0.88 (12th)** | **0.89 (5th)** | **0.58 (10th)** | **0.75 (16th)** |
| **Moirai-2** | **0.94 (13th)** | **1.10 (16th)** | **0.49 (1st)** | **0.80 (14th)** |
| OHT_JHU-nbxd | 0.95 (14th) | 1.07 (14th) | 0.27 (17th) | 0.60 (17th) |
| **CovidHub-baseline** | **1.00 (15th)** | **1.00 (10th)** | **0.67 (16th)** | **0.93 (8th)** |
| UM-DeepOutbreak | 1.06 (16th) | 1.09 (15th) | 0.67 (15th) | 0.92 (4th) |
| **Lag-Llama** | **1.58 (17th)** | **1.80 (17th)** | **0.58 (11th)** | **0.92 (5th)** |

**Table S4. Aggregated forecasting performance of all CovidHub models and TSFMs (N = 17), season 2025/26.** Columns report relative log WIS (Rel. log WIS), relative log AE (Rel. log AE), and 50% and 90% prediction interval coverage. Rankings in parentheses refer to all models in the challenge; for coverage, rank 1 corresponds to the model closest to the nominal level (0.50 and 0.90, respectively). In bold are the TSFMs, the ensemble, and the baseline. Models are sorted by Rel. log WIS. TSFMs, time series foundation models; WIS, weighted interval score; AE, absolute error.

Figures S1 and S2 present the subnational performance analysis in terms of relative log WIS and AE, respectively, for FluSight (A) and CovidHub (B). Models are sorted by ascending median value, and the values of the pairwise comparison score are directly reported. Values below 1 indicate better performance relative to the hub-baseline model.

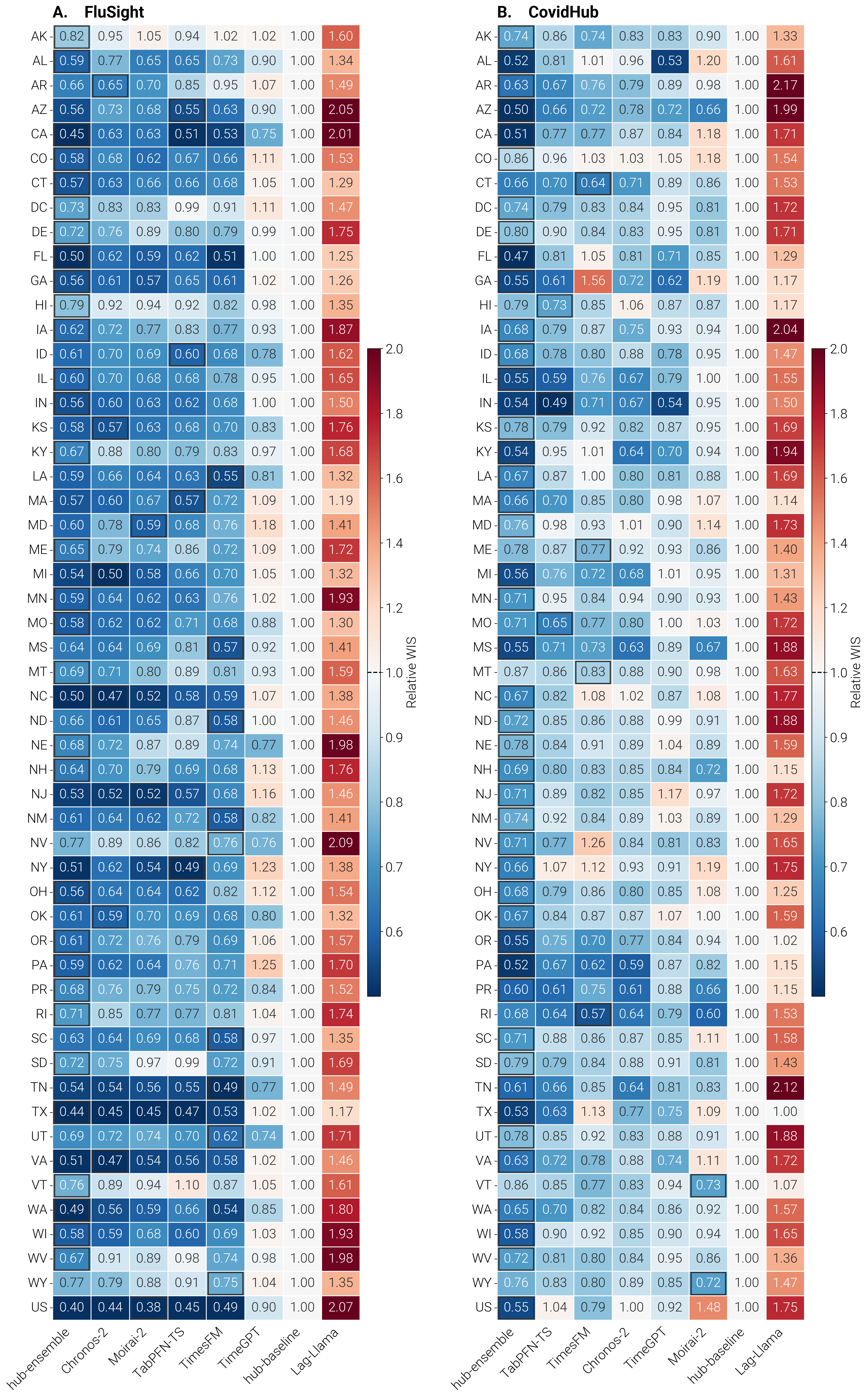

**Figure S1. TSFMs performance across locations, 2025/26 season. Relative log WIS of TSFMs and hub models across U.S. states and the national level for FluSight (A) and CovidHub (B).** Models are ordered by increasing median, and the best-performing model for each state is highlighted with a black border. The baseline equals 1 by construction. TSFMs, time series foundation models; WIS, weighted interval score.

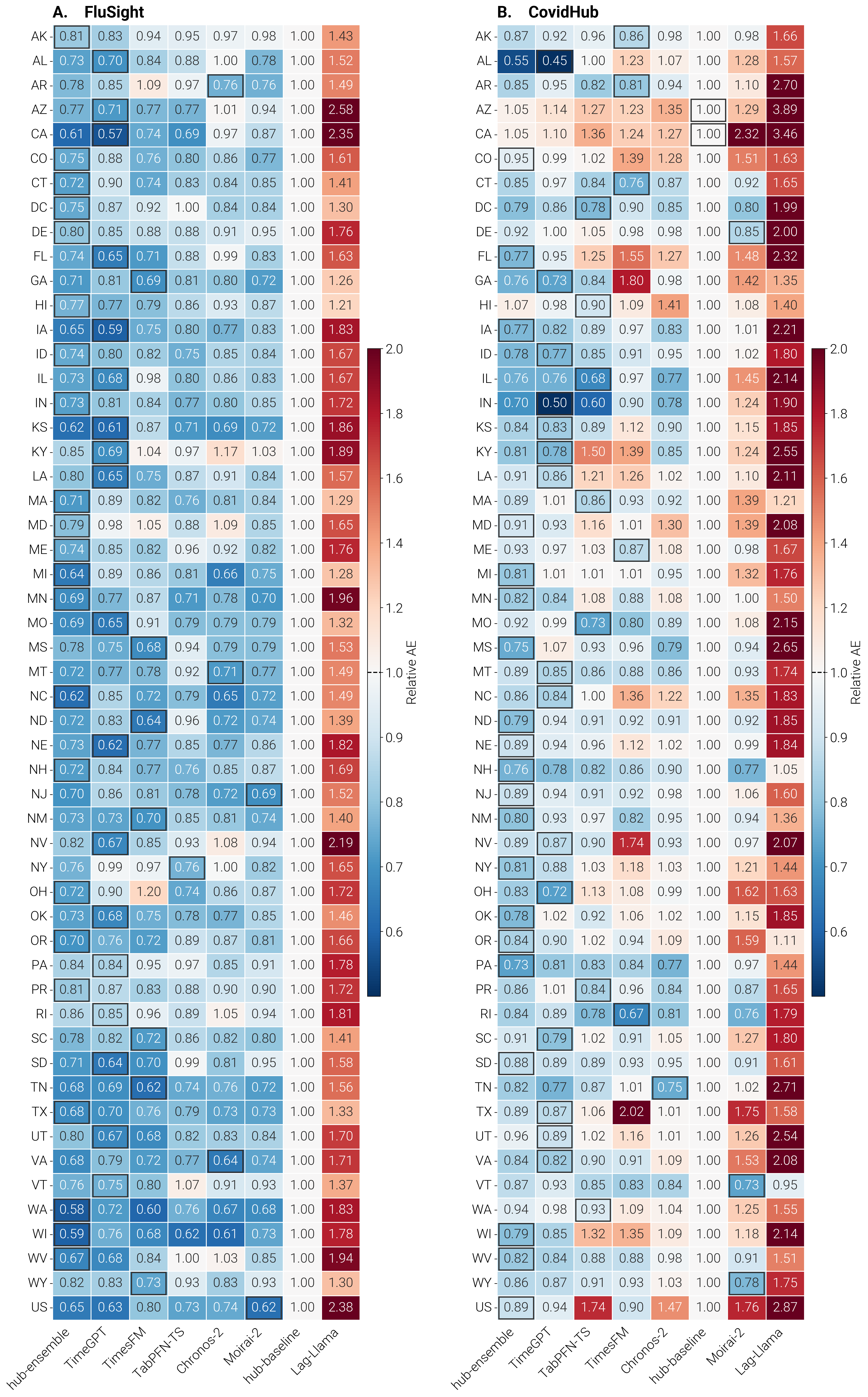

**Figure S2. TSFMs performance across locations, 2025/26 season. Relative log AE of TSFMs and hub models across U.S. states and the national level for FluSight (A) and CovidHub (B).** Models are ordered by increasing median, and the best-performing model for each state is highlighted with a black border. The baseline equals 1 by construction. TSFMs, time series foundation models; AE, absolute error.

**S3 Retrospective evaluation of TSFMs against multi-model ensembles and individual models during the 2024/25 season**

Here, we retrospectively evaluate TSFM’s performance considering the 2024/25 season. Again, we use as case studies both influenza and COVID-19. During the 2024/25 season, 38 models were eligible for evaluation for FluSight and 10 models for CovidHub; we refer to Sec. S1 for more details on individual models contributed to the forecasting challenge, eligibility for evaluation, and model types. We use versioned data that would have been available at the time each forecast was made, over 25 forecasting rounds from November 30, 2024 through May 24, 2025. We repeat all the analyses conducted for the 2025/26 season. First, we evaluate aggregated performance across U.S. locations (both U.S. national, and 52 jurisdictions) of all individual models, the ensemble, the baseline models, and the six evaluated TSFMs using the pairwise comparison approach detailed in the manuscript in the Methods, to account for the fact that not all models submit predictions for the same set of tasks (i.e., not all locations, not all weeks, not all horizons). Tables S5 and S6 report the aggregated forecasting performance for FluSight and CovidHub, respectively. TSFMs are highlighted in bold, as well as the hub ensemble and baseline models. Models are sorted by Rel. log WIS, and in parentheses are reported the ranking of each model relative to the full set of contributing models.

For FluSight, 44 models were evaluated in total, including the six TSFMs. All TSFMs show a relative log WIS and AE below 1, indicating better performance than the baseline, with the exception of Lag-Llama and TimeGPT for relative log WIS and only Lag-Llama for relative log AE. Chronos-2 ranks in the top tertile for relative log WIS (10th), 50% coverage (14th), and 90% coverage level (1st), and in the middle tertile for relative log AE (19th). TabPFN-TS placed in the middle tertile for all metrics: relative log WIS (17th), relative log AE (21st), 50% coverage (27th), 90% coverage (24th). TimesFM placed in the middle tertile for relative log WIS (20th), relative log AE (17th), and 90% coverage (30th), and in the bottom tertile for 50% coverage (32nd). Moirai-2 placed in the middle tertile for relative log WIS (24th), relative log AE (23rd), and 50% coverage (29th), and in the bottom tertile for 90% coverage (36th). TimeGPT ranked in the bottom tertile for relative log WIS (37th) and in the middle tertile for relative log AE (24th), 50% coverage (23rd), and 90% coverage (26th), confirming the cross-metric inconsistency observed in the 2025/26 season. Lag-Llama ranked last among all models for both relative log WIS (44th) and relative log AE (44th), placing in the middle tertile for 50% coverage (20th) but in the bottom tertile for 90% coverage (37th). In comparison, the FluSight ensemble ranked in the top tertile across all metrics (3rd for relative log WIS, 2nd for relative log AE, 5th for 50% coverage, and 9th for 90% coverage).

For the CovidHub, 16 models were evaluated in total, including the six TSFMs. TabPFN-TS placed in the top third for relative log WIS (5th), relative log AE (3rd), and 50% coverage (4th), and in the middle tertile for 90% coverage (9th). TimesFM ranked in the top third for relative log WIS (6th) and 50% coverage (1st), and in the middle tertile for relative log AE (8th) and 90% coverage (10th). Moirai-2 ranked in the middle tertile for relative log WIS (10th), relative log AE (10th), and 50% coverage (8th), and in the bottom tertile for 90% coverage (13th). Chronos-2 placed in the middle tertile for relative log WIS (7th) and relative log AE (9th), in the top tertile for 90% coverage (3rd), and in the bottom tertile for 50% coverage (13th). TimeGPT again exhibited cross-metric inconsistency, ranking in the top tertile for relative log AE (2nd) and 50% coverage (6th), but in the middle tertile for relative log WIS (9th) and bottom for 90% coverage (16th). Lag-Llama ranked last among all models for both relative log WIS (16th) and relative log AE (16th), with 90% coverage in the middle tertile (8th) and 50% coverage in the bottom tertile (12th). In comparison, the CovidHub ensemble ranked in the top tertile for all metrics: relative log WIS (1st), relative log AE (1st), 50% coverage (5th), and 90% coverage (2nd). Overall, these results confirm the robustness of the multi-model ensemble across both diseases and metrics and highlight TSFMs heterogeneity across diseases and metrics.

| **Model** | **Rel. log WIS** | **Rel. log AE** | **50% Cov.** | **90% Cov.** |
| --- | --- | --- | --- | --- |
| PSI-PROF_beta | 0.61 (1st) | 0.72 (3rd) | 0.57 (17th) | 0.85 (3rd) |
| UMass-flusion | 0.63 (2nd) | 0.74 (6th) | 0.40 (24th) | 0.76 (12th) |
| **FluSight-ensemble** | **0.64 (3rd)** | **0.71 (2nd)** | **0.52 (5th)** | **0.78 (9th)** |
| PSI-PROF | 0.64 (4th) | 0.80 (11th) | 0.54 (10th) | 0.85 (4th) |
| UGA_flucast-Scenariocast | 0.64 (5th) | 0.79 (9th) | 0.47 (8th) | 0.84 (6th) |
| ISU_NiemiLab-GPE | 0.65 (6th) | 0.70 (1st) | 0.46 (11th) | 0.72 (22nd) |
| CU-ensemble | 0.65 (7th) | 0.74 (5th) | 0.51 (1st) | 0.78 (8th) |
| CMU-TimeSeries | 0.66 (8th) | 0.78 (7th) | 0.53 (6th) | 0.86 (2nd) |
| NEU_ISI-AdaptiveEnsemble | 0.66 (9th) | 0.73 (4th) | 0.29 (38th) | 0.61 (38th) |
| **Chronos-2** | **0.67 (10th)** | **0.82 (19th)** | **0.44 (14th)** | **0.87 (1st)** |
| UGA_flucast-INFLAenza | 0.67 (11th) | 0.82 (20th) | 0.41 (22nd) | 0.77 (10th) |
| OHT_JHU-nbxd | 0.68 (12th) | 0.79 (8th) | 0.39 (25th) | 0.72 (21st) |
| NIH-Flu_ARIMA | 0.68 (13th) | 0.80 (13th) | 0.25 (40th) | 0.59 (39th) |
| LosAlamos_NAU-CModel_Flu | 0.69 (14th) | 0.87 (26th) | 0.32 (36th) | 0.75 (15th) |
| CEPH-Rtrend_fluH | 0.71 (15th) | 0.82 (16th) | 0.43 (16th) | 0.75 (18th) |
| UGA_flucast-Copycat | 0.72 (16th) | 0.84 (22nd) | 0.39 (28th) | 0.76 (13th) |
| **TabPFN-TS** | **0.73 (17th)** | **0.82 (21st)** | **0.39 (27th)** | **0.69 (24th)** |
| NEU_ISI-FluBcast | 0.73 (18th) | 0.80 (12th) | 0.26 (39th) | 0.50 (40th) |
| MIGHTE-Joint | 0.77 (19th) | 0.81 (15th) | 0.38 (30th) | 0.62 (34th) |
| **TimesFM** | **0.77 (20th)** | **0.82 (17th)** | **0.37 (32nd)** | **0.64 (30th)** |
| MIGHTE-Nsemble | 0.79 (21st) | 0.82 (18th) | 0.37 (31st) | 0.62 (35th) |
| VTSanghani-PRIME | 0.79 (22nd) | 0.79 (10th) | 0.59 (21st) | 0.75 (16th) |
| JHU_CSSE-CSSE_Ensemble | 0.80 (23rd) | 0.86 (25th) | 0.51 (3rd) | 0.77 (11th) |
| **Moirai-2** | **0.82 (24th)** | **0.85 (23rd)** | **0.38 (29th)** | **0.61 (36th)** |
| UNC_IDD-InfluPaint | 0.82 (25th) | 0.81 (14th) | 0.12 (43rd) | 0.37 (42nd) |
| UMass-AR2 | 0.83 (26th) | 0.93 (29th) | 0.42 (19th) | 0.74 (19th) |
| MOBS-GLEAM_FLUH | 0.87 (27th) | 0.89 (27th) | 0.19 (41st) | 0.46 (41st) |
| LUcompUncertLab-chimera | 0.88 (28th) | 0.95 (31st) | 0.35 (34th) | 0.66 (28th) |
| UMass-trends_ensemble | 0.89 (29th) | 0.89 (28th) | 0.42 (18th) | 0.64 (31st) |
| Gatech-ensemble_point | 0.90 (30th) | 1.03 (36th) | 0.34 (35th) | 0.68 (27th) |
| UVAFluX-Ensemble | 0.94 (31st) | 1.01 (34th) | 0.56 (13th) | 0.75 (14th) |
| SigSci-TSENS | 0.97 (32nd) | 0.94 (30th) | 0.45 (12th) | 0.64 (32nd) |
| **FluSight-baseline** | **1.00 (33rd)** | **1.00 (33rd)** | **0.32 (37th)** | **0.63 (33rd)** |
| JHUAPL-DMD | 1.00 (34th) | 1.02 (35th) | 0.49 (2nd) | 0.78 (7th) |
| fjordhest-ensemble | 1.07 (35th) | 1.17 (40th) | 0.44 (15th) | 0.73 (20th) |
| UVAFluX-CESGCN | 1.07 (36th) | 0.98 (32nd) | 0.15 (42nd) | 0.35 (43rd) |
| **TimeGPT** | **1.09 (37th)** | **0.85 (24th)** | **0.59 (23rd)** | **0.68 (26th)** |
| Gatech-ensemble_prob | 1.15 (38th) | 1.21 (41st) | 0.36 (33rd) | 0.65 (29th) |
| CMU-climate_baseline | 1.18 (39th) | 1.34 (42nd) | 0.47 (7th) | 0.85 (5th) |
| UGA_CEID-Walk | 1.19 (40th) | 1.16 (39th) | 0.48 (4th) | 0.70 (23rd) |
| UI_CompEpi-EpiGen | 1.24 (41st) | 1.13 (38th) | 0.54 (9th) | 0.69 (25th) |
| UM-DeepOutbreak | 1.26 (42nd) | 1.03 (37th) | 0.61 (26th) | 0.75 (17th) |
| Stevens-ILIForecast | 1.58 (43rd) | 1.44 (43rd) | 0.05 (44th) | 0.11 (44th) |
| **Lag-Llama** | **1.82 (44th)** | **1.80 (44th)** | **0.42 (20th)** | **0.61 (37th)** |

**Table S5. Aggregated forecasting performance of all FluSight and TSFMs (N = 44), season 2024/25.** Columns report relative log WIS (Rel. log WIS), relative log AE (Rel. log AE), and 50% and 90% prediction interval coverage. Rankings in parentheses refer to all models in the challenge; for coverage, rank 1 corresponds to the model closest to the nominal level (0.50 and 0.90, respectively). In bold are the TSFM models, the ensemble, and the baseline. Models are sorted by Rel. log WIS. TSFMs, time series foundation models; WIS, weighted interval score; AE, absolute error.

| **Model** | **Rel. log WIS** | **Rel. log AE** | **50% Cov.** | **90% Cov.** |
| --- | --- | --- | --- | --- |
| **CovidHub-ensemble** | **0.69 (1st)** | **0.89 (1st)** | **0.55 (5th)** | **0.89 (2nd)** |
| CMU-TimeSeries | 0.75 (2nd) | 0.96 (4th) | 0.52 (3rd) | 0.89 (4th) |
| UMass-ar6_pooled | 0.78 (3rd) | 1.00 (6th) | 0.56 (9th) | 0.90 (1st) |
| UMass-gbqr | 0.80 (4th) | 0.98 (5th) | 0.44 (10th) | 0.81 (12th) |
| **TabPFN-TS** | **0.84 (5th)** | **0.95 (3rd)** | **0.54 (4th)** | **0.83 (9th)** |
| **TimesFM** | **0.88 (6th)** | **1.06 (8th)** | **0.49 (1st)** | **0.83 (10th)** |
| **Chronos-2** | **0.89 (7th)** | **1.07 (9th)** | **0.37 (13th)** | **0.89 (3rd)** |
| CEPH-Rtrend_covid | 0.91 (8th) | 1.22 (13th) | 0.45 (7th) | 0.81 (11th) |
| **TimeGPT** | **0.92 (9th)** | **0.91 (2nd)** | **0.45 (6th)** | **0.63 (16th)** |
| **Moirai-2** | **0.92 (10th)** | **1.09 (10th)** | **0.44 (8th)** | **0.78 (13th)** |
| OHT_JHU-nbxd | 0.94 (11th) | 1.12 (11th) | 0.32 (15th) | 0.66 (14th) |
| **CovidHub-baseline** | **1.00 (12th)** | **1.00 (7th)** | **0.61 (11th)** | **0.85 (7th)** |
| MOBS-GLEAM_COVID | 1.07 (13th) | 1.31 (14th) | 0.30 (16th) | 0.65 (15th) |
| CMU-climate_baseline | 1.21 (14th) | 1.69 (15th) | 0.52 (2nd) | 0.94 (5th) |
| UM-DeepOutbreak | 1.24 (15th) | 1.14 (12th) | 0.65 (14th) | 0.94 (6th) |
| **Lag-Llama** | **1.75 (16th)** | **2.00 (16th)** | **0.62 (12th)** | **0.95 (8th)** |

**Table S6. Aggregated forecasting performance of all CovidHub and TSFMs (N = 16), season 2024/25.** Columns report relative log WIS (Rel. log WIS), relative log AE (Rel. log AE), and 50% and 90% prediction interval coverage. Rankings in parentheses refer to all models in the challenge; for coverage, rank 1 corresponds to the model closest to the nominal level (0.50 and 0.90, respectively). In bold are the TSFM models, the ensemble, and the baseline. Models are sorted by Rel. log WIS. TSFMs, time series foundation models; WIS, weighted interval score; AE, absolute error.

Fig. S3 presents the location-level performance analysis for the 2024/25 season, showing TSFM performance across individual U.S. jurisdictions. Panels A and C show the distribution of relative log WIS for FluSight and CovidHub, respectively, for the TSFMs, hub ensemble, and baseline models, sorted by ascending median value. The corresponding figure for relative log AE is reported in panels B and D. Each dot represents the relative log WIS (or AE) score for a specific U.S. jurisdiction; the empty square marker indicates the score for U.S. national forecasts, and the median across jurisdictions for each model is highlighted with a diamond-filled marker. The lateral bars report the number of jurisdictions for which each model ranks first among these models.

Focusing on FluSight (Figs. S3A, S3B), models fall into three broad groups. Similar to the 2025/26 season, Chronos-2, TimesFM, TabPFN-TS, and Moirai-2 consistently outperform the baseline, with median relative log WIS and AE below 1 in the majority of locations (more than 80%). Lag-Llama systematically underperforms, with both metrics above 1 across all locations. TimeGPT exhibits a mixed pattern again: it outperforms the baseline in less than 30% of locations for relative log WIS, but in nearly all locations for relative log AE, suggesting stronger point-forecast accuracy than interval sharpness and calibration. The FluSight ensemble achieves the lowest median relative log WIS and median relative log AE across all locations. Among the six TSFMs, the ensemble, and the baseline, the FluSight ensemble ranks first for relative log WIS in the most jurisdictions (33 out of 52), with Chronos-2 second (11 states). For relative log AE, the ensemble again leads ranking first in 39 states.

Considering the CovidHub (Fig. S3C, S3D), we observe broadly similar patterns across models. The ensemble achieves the lowest median relative log WIS and AE and outperforms the baseline in nearly all locations (51 of 53 for relative log WIS and 45 of 53 for relative log AE). TabPFN-TS outperforms the baseline on both metrics, while TimesFM and Chronos-2 outperform it only for Rel. log WIS (median relative log AE of 1.06 and 1.07), with median values closer to 1 than FluSight’s. Moirai-2 shows more pronounced borderline performance, with median values closer to or slightly higher than 1 considering both metrics. TimeGPT again exhibits the same cross-metric inconsistency observed for FluSight, with lower outperformance rates for relative log WIS than for relative log AE. Lag-Llama continues to underperform systematically, falling below the baseline in almost all locations for both relative log WIS and AE. Among the six TSFMs, the ensemble and the baseline, the ensemble ranks first for both relative log WIS and AE in the greatest number of jurisdictions (38 out of 52 and 23 out of 52, respectively). For relative log AE, TimeGPT follows the ensemble ranking first in 13 states, consistent with its stronger point forecast accuracy relative to interval calibration.

Figures S4 and S5 show the relative log WIS and AE for each location, with panel A referring to FluSight and panel B to CovidHub. A black border marks the best-performing model (lowest value) per location among the six TSFMs, the hub ensemble, and the baseline.

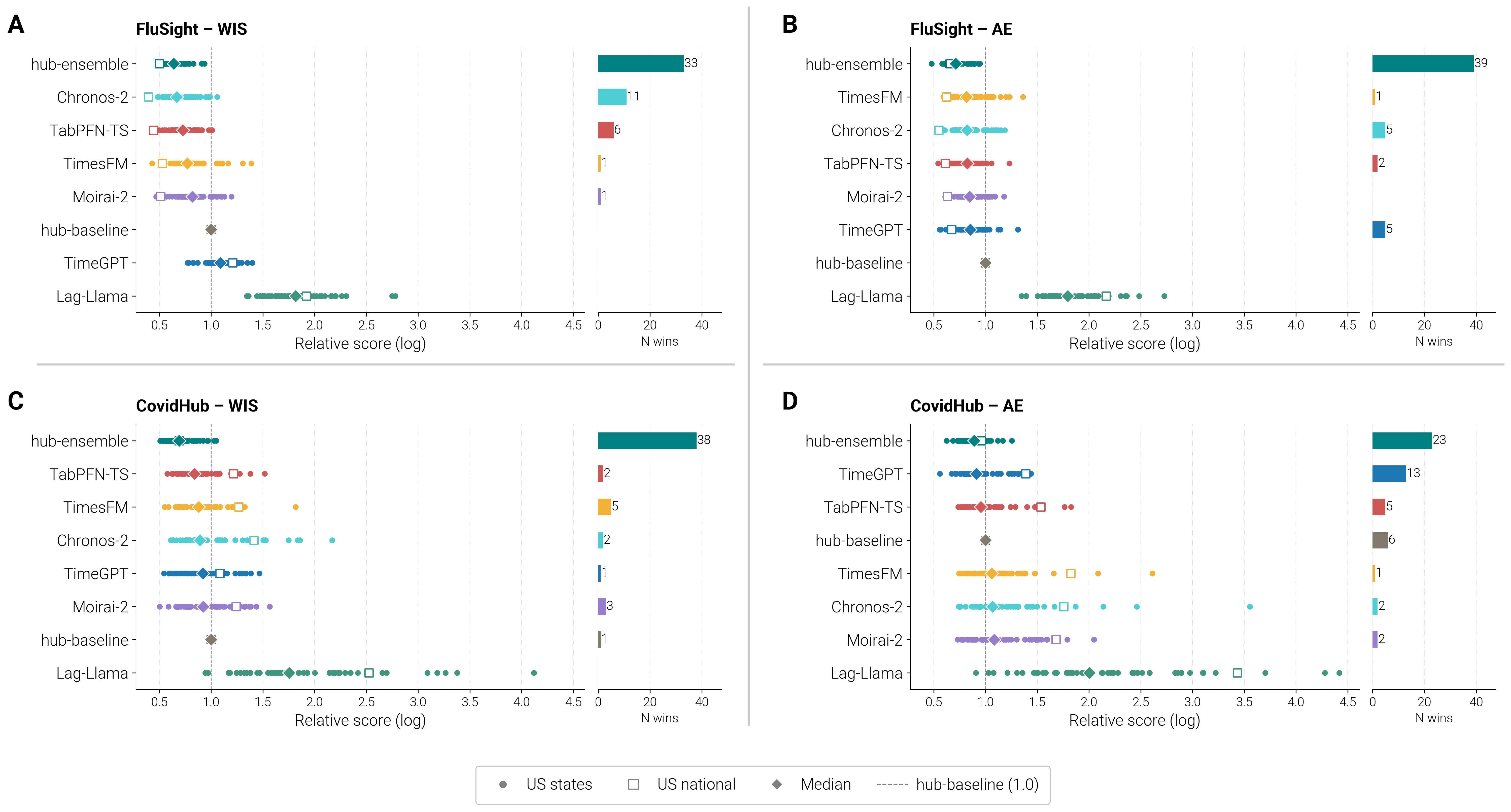

**Figure S3. TSFMs performance across locations, 2024/25 season. Relative log WIS and AE of TSFMs and hub models across U.S. states and the national level for FluSight (A, B) and CovidHub (C, D).** Each dot represents a state, the square marker represents the U.S. national, and the diamond indicates the median across all locations. The bars on the right (N wins) represent the number of states in which the model ranks first (i.e., lowest score value). Models are ordered by increasing median. TSFMs, time series foundation models; WIS, weighted interval score; AE, absolute error.

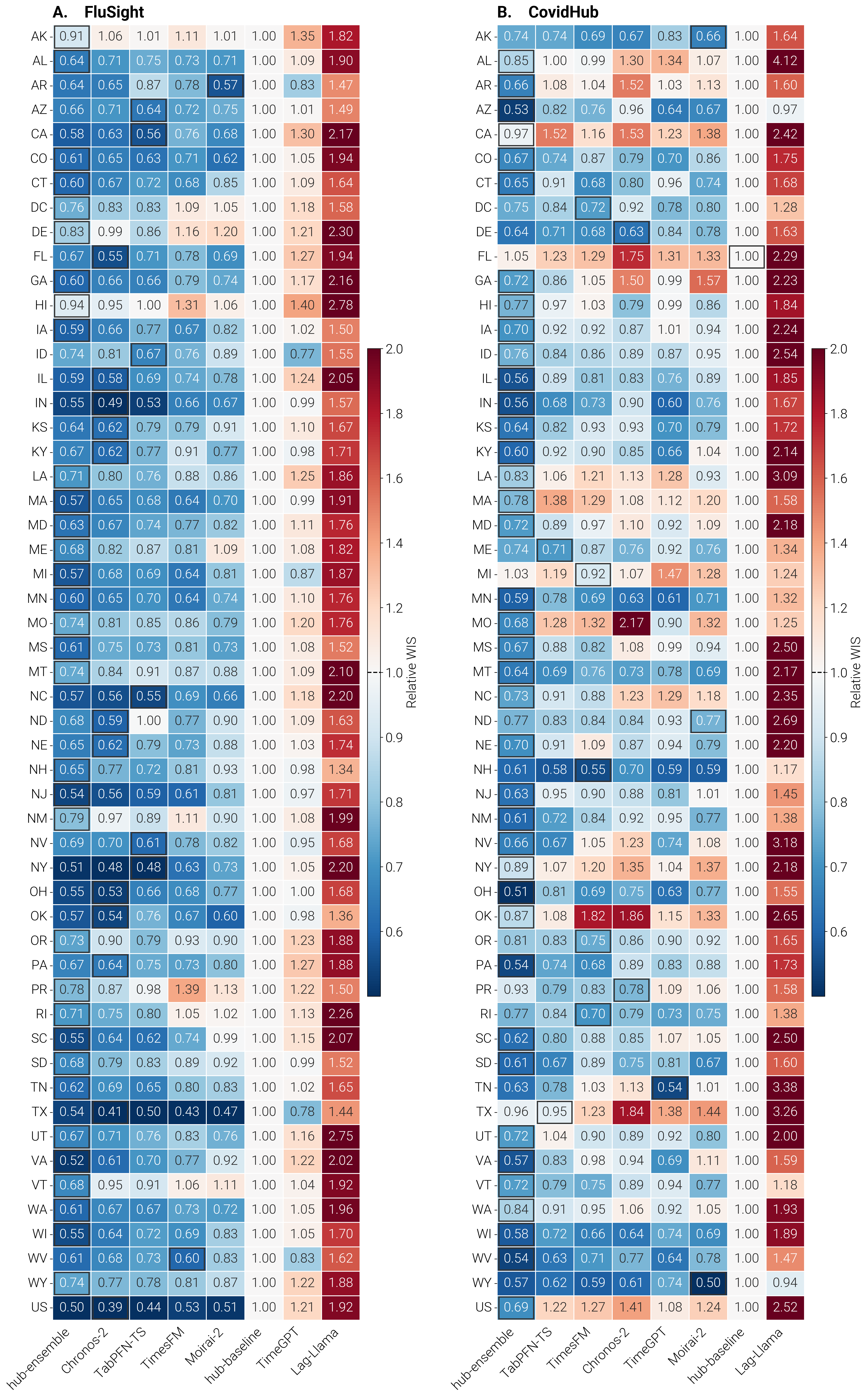

**Figure S4. TSFMs performance across locations, 2024/25 season. Relative log WIS of TSFMs and hub models across U.S. states and the national level for FluSight (A) and CovidHub (B).** Models are ordered by increasing median, and the best-performing model for each state is highlighted with a black border. The baseline equals 1 by construction. TSFMs, time series foundation models; WIS, weighted interval score.

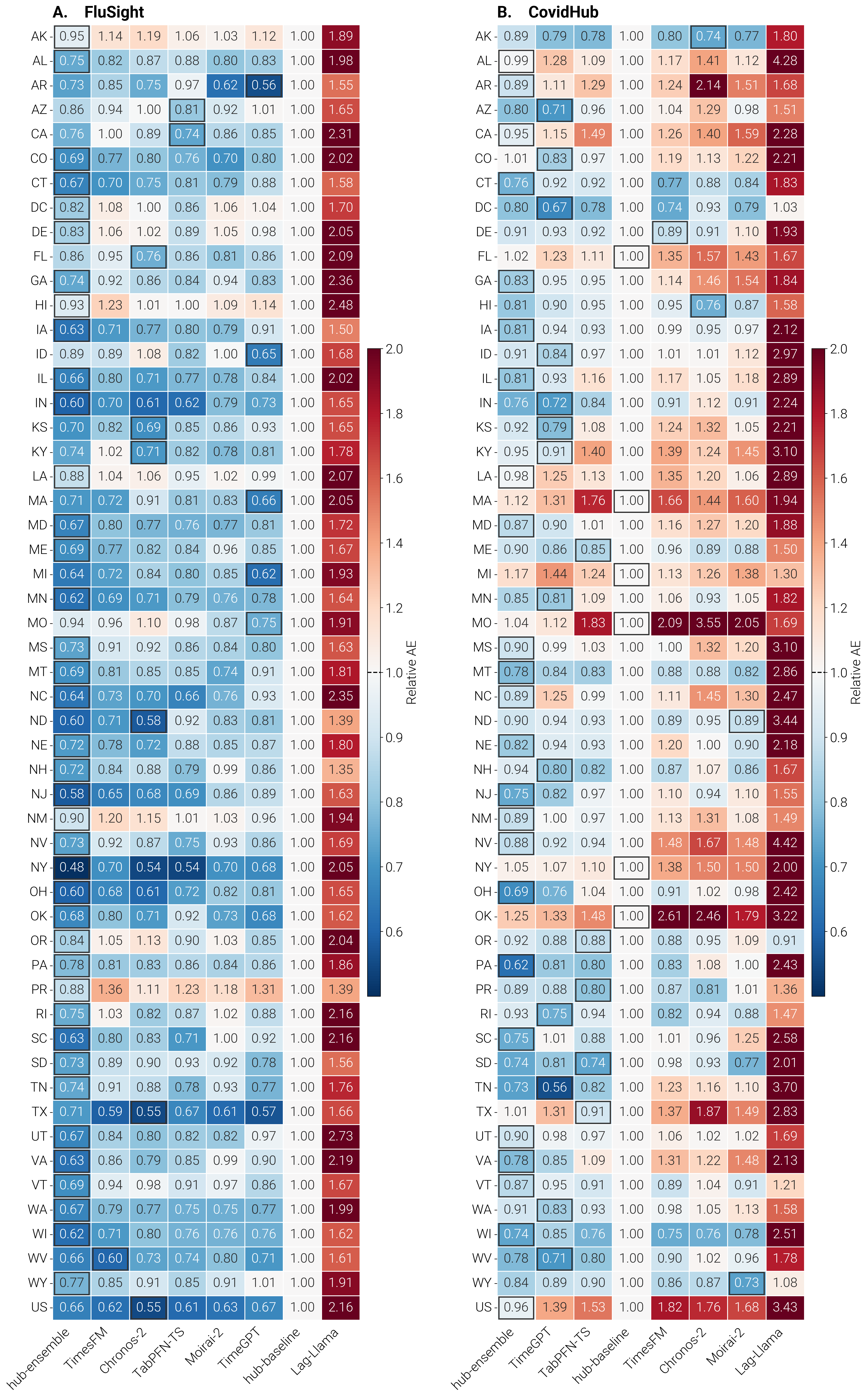

**Figure S5. TSFMs performance across locations, 2024/25 season. Relative log AE of TSFMs and hub models across U.S. states and the national level for FluSight (A) and CovidHub (B).** Models are ordered by increasing median, and the best-performing model for each state is highlighted with a black border. The baseline equals 1 by construction. TSFMs, time series foundation models; AE, absolute error.

We explore forecasting performance consistency for the 2024/25 season by calculating, for each model, the percentage of forecasts in which it ranked in the top and bottom 30% of models. This analysis ranks models directly on their log WIS (or log AE) within each forecast instance, defined by reference date, location, and horizon. We then build the performance distribution map, with each point corresponding to a model and its coordinates are the frequencies of top (y-axis) and bottom (x-axis) 30% placements. As described in the main text, the median values along both axes define four quadrants that reflect consistency performance: Top performers (low proportion of bottom-ranked forecasts and high proportion of top-ranked forecasts); Consistent performers (low frequency in both axes); Inconsistent performers (high frequency in both axes); Frequent low performers (high bottom-ranked forecasts and low top-ranked forecasts). Figure S6 shows these maps for FluSight (top row) and CovidHub (bottom row), for log WIS (panels A, C) and log AE (panels B, D). Colored circles denote TSFMs, grey circles other individual models, a square the baseline, and a diamond the ensemble. The overall structure shows what we observed for 2025/26: models cluster along the anti-diagonal, and few models fall in the top-right corner, confirming that high frequencies at both extremes, reflecting high inconsistency, is uncommon. Relative to 2025/26, however, TSFMs are more concentrated in the central region of the plot for both metrics, suggesting moderate and mixed performance rather than clear dominance or persistent underperformance. The ensemble achieves a proportion of top-30% ranked forecasts comparable to the TSFMs for FluSight and higher than all TSFMs for CovidHub log WIS, while almost never falling into the bottom 30%, underscoring its reliability across rounds.

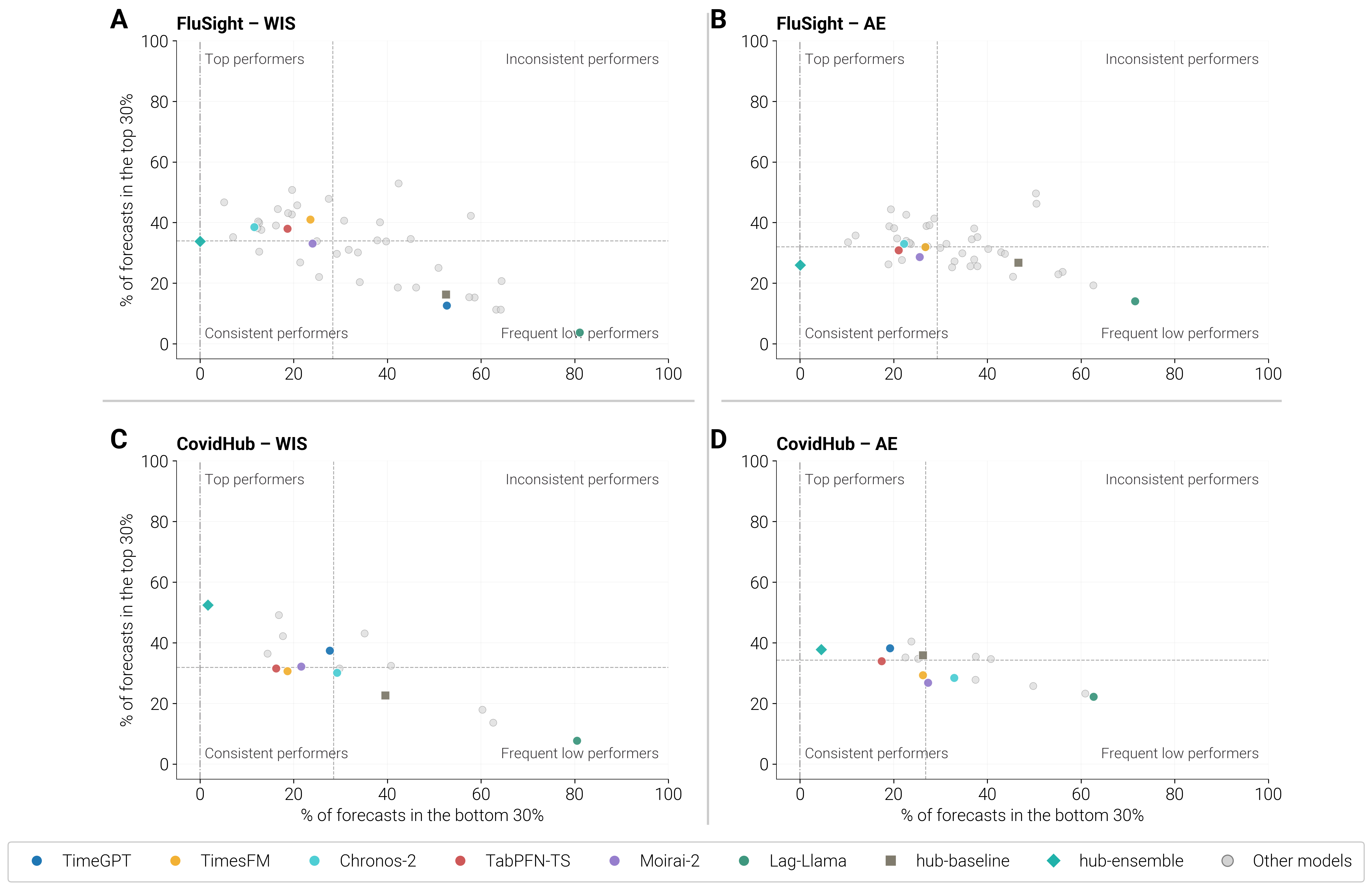

**Figure S6. Performance distribution of individual models and the ensemble for FluSight and CovidHub across the 2024/25 season based on log WIS (panels A, C) and log AE (panels B, D) rankings.** The x-axis shows the percentage of forecasts in which a model ranked in the bottom 30%, while the y-axis shows the percentage of forecasts in which the model ranked in the top 30%. Each colored circle represents a specific TSFM, the diamond represents the ensemble model, the square represents the baseline model, and grey circles represent individual models. Quadrants are defined using the median values of both axes across all models, defining the following categories: Top performers (low bottom-30% frequency, high top-30% frequency), Consistent performers (low frequency in both bottom-30% and top-30%), Inconsistent performers (high frequency in both bottom-30% and top-30%), and Frequent low performers (high bottom-30% frequency, low top-30% frequency). TSFM, time series foundation model; WIS, weighted interval score; AE, absolute error.

Finally, we evaluate performance stratified by epidemic phase, classifying each week into one of five phases based on the week-to-week difference in hospital admission rates: large decrease, decrease, stable, increase, large increase. For each forecast instance, defined by reference date, location, and horizon, we compute the model's log WIS (or log AE) relative to that of the baseline. Figure S7 presents these results, which reveal a notably different picture from the 2025/26 season and a more mixed overall pattern. Considering FluSight, during the large decrease and decrease trends, most models perform better relative to the stable, increase and large increase trends, as indicated by the leftward shift of the blue and light blue dots relative to the red ones. This stands in contrast to the 2025/26 season, where performance improved during the stable and increase trends. More mixed results are observed in the CovidHub, especially when evaluated using the log WIS ratio, while considering the log AE ratio we observe that forecasts during the decrease trends perform worse than the increase ones. Lag-Llama again shows markedly lower performance across all phases compared to other models. The ensemble ranks among the top three positions in most epidemic phases, with the exceptions of the decrease phase for FluSight log WIS (4th) and the large increase phase for CovidHub (6th for both log WIS and log AE).

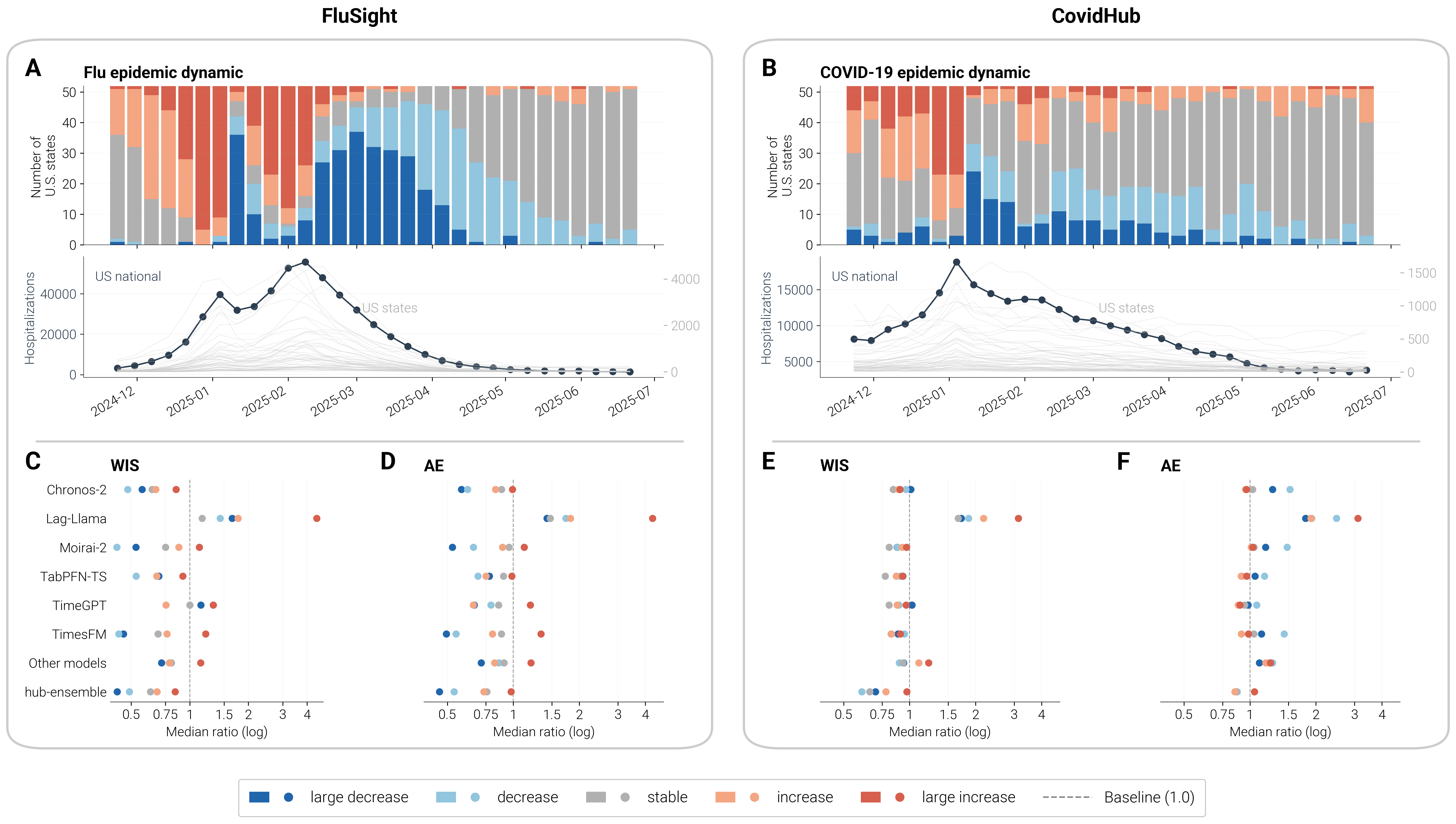

**Figure S7. Performance stratified by epidemic phases for FluSight (left panel) and CovidHub (right panel) across the 2024/25 season.** Panels A and B show the number of states assigned to each phase per week (top row) and the U.S. national surveillance curve alongside the individual state-level curves in light grey (bottom row). Panels C–F show performance stratified by epidemic phases based on log WIS (C, E) and log AE (D, F) ratios compared to the baseline. Each dot represents the median score ratio for one epidemic phase: blue for large decrease, light blue for decrease, grey for stable, orange for increase, and red for large increase. Values to the left of the dashed line (ratio < 1) indicate better performance than the baseline. The x-axis is on a log scale. TSFMs are shown in alphabetical order; for reference, the hub ensemble and the average across all individual contributing models are also reported. TSFMs, time series foundation models; WIS, weighted interval score; AE, absolute error.

**S4 TSFMs pre-processing and post-processing corrections**

As described in the main text, several pre- and post-processing steps were applied to ensure consistent inputs and outputs across the six TSFMs, so that predictions conformed to the output format required by the collaborative hubs. Table S7 summarizes, for each model, which of the following steps were applied: i) gap-filling of missing input values via linear interpolation, required for models needing a complete input sequence; ii) post-hoc correction of non-monotonic quantile outputs, in which flagged quantiles were replaced via linear interpolation between the nearest valid neighbors; iii) interpolation of missing quantile levels, applied to models that do not natively produce all 23 required quantile levels; and iv) clipping of negative quantile values to zero.

| **Model** | **Gap-free input sequence required** | **Monotonicity**  **quantiles violation** | **Incomplete quantiles sequence (23 quantiles)** | **Negative output values** |
| --- | --- | --- | --- | --- |
| Chronos-2 | × | ✓ | × | ✓ |
| Lag-Llama | × | × | × | ✓ |
| Moirai-2 | ✓ | ✓ | ✓ (9 quantiles) | ✓ |
| TabPFN-TS | ✓ | × | ✓ (9 quantiles) | ✓ |
| TimeGPT | ✓ | × | × | ✓ |
| TimesFM | × | × | ✓ (9 quantiles) | × |

**Table S7. Pre- and post-processing steps applied to each TSFM to ensure consistent input and output formats.** Checkmarks (✓) indicate that a given correction step was applied to the corresponding model, and crosses (×) indicate that it was not required. Columns correspond to: gap-filling of missing input values via linear interpolation, required for models needing a complete, gap-free input sequence; post-hoc correction of non-monotonic quantile outputs, in which flagged quantiles were replaced via linear interpolation between the nearest valid neighbors; interpolation of missing quantile levels, applied to models that do not natively produce all 23 required quantile levels (the number of natively produced quantile levels is indicated in parentheses); and clipping of negative quantile output values to zero. TSFM, time series foundation model.

Fig. S8 shows the percentage of negative quantile values produced by each TSFM, broken down by reference date (A, C) and by location (B, D), during the 2025/26 season, for FluSight and CovidHub. Considering FluSight, Lag-Llama and TimeGPT show the highest percentages of negative values, for both reference dates and locations, and particularly in the second half of the period, while Chronos-2, Moirai-2, and TabPFN-TS show comparatively low or negligible percentages throughout. Only TimesFM produced no negative quantile values. For CovidHub, we observe a broadly similar pattern, with Lag-Llama consistently producing the highest percentage of negative values across both reference dates and locations, and TimesFM the only model producing no negative quantile values. In this case, most TSFMs show negative values from end of March, 2026, onwards. Fig. S9 presents the same visualization for the 2024/25 season, with overall consistent results, except for negative values by reference date for the CovidHub, in which the only model affected by relatively high proportion of negative values is Lag-Llama, with other TSFMs less affected compared to 2025/26 season.

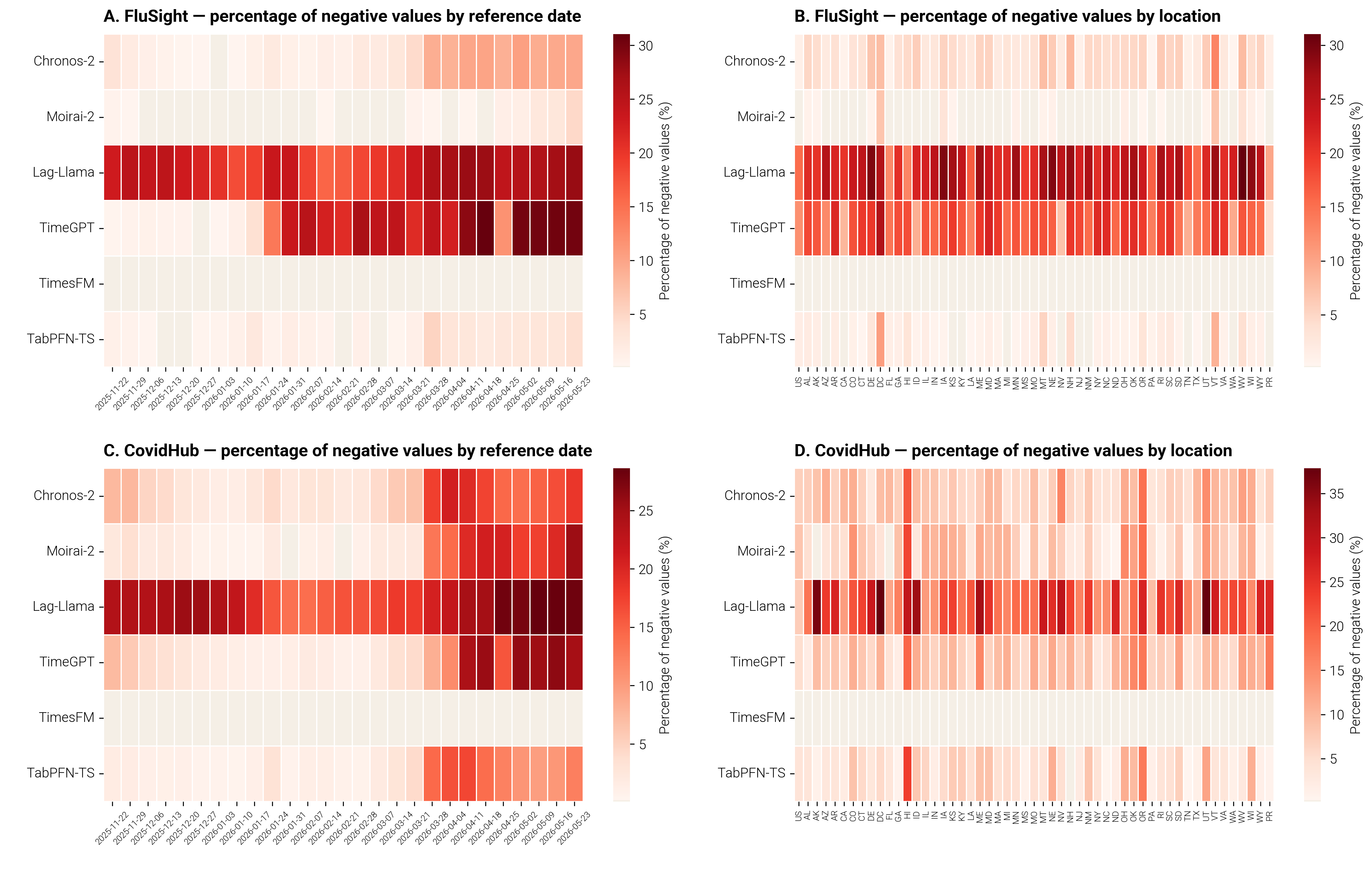

**Figure S8. Percentage of negative quantile values produced by each TSFM for FluSight and CovidHub, by reference date (A, C) and by location (B, D), season 2025/26.** For each model, colored cells indicate the percentage of quantile forecasts with negative predicted values, computed separately across all reference dates (A, C) and all locations (B, D), before values were clipped to zero. Cells shown in ivory correspond to a value of 0%. TSFM, time series foundation model.

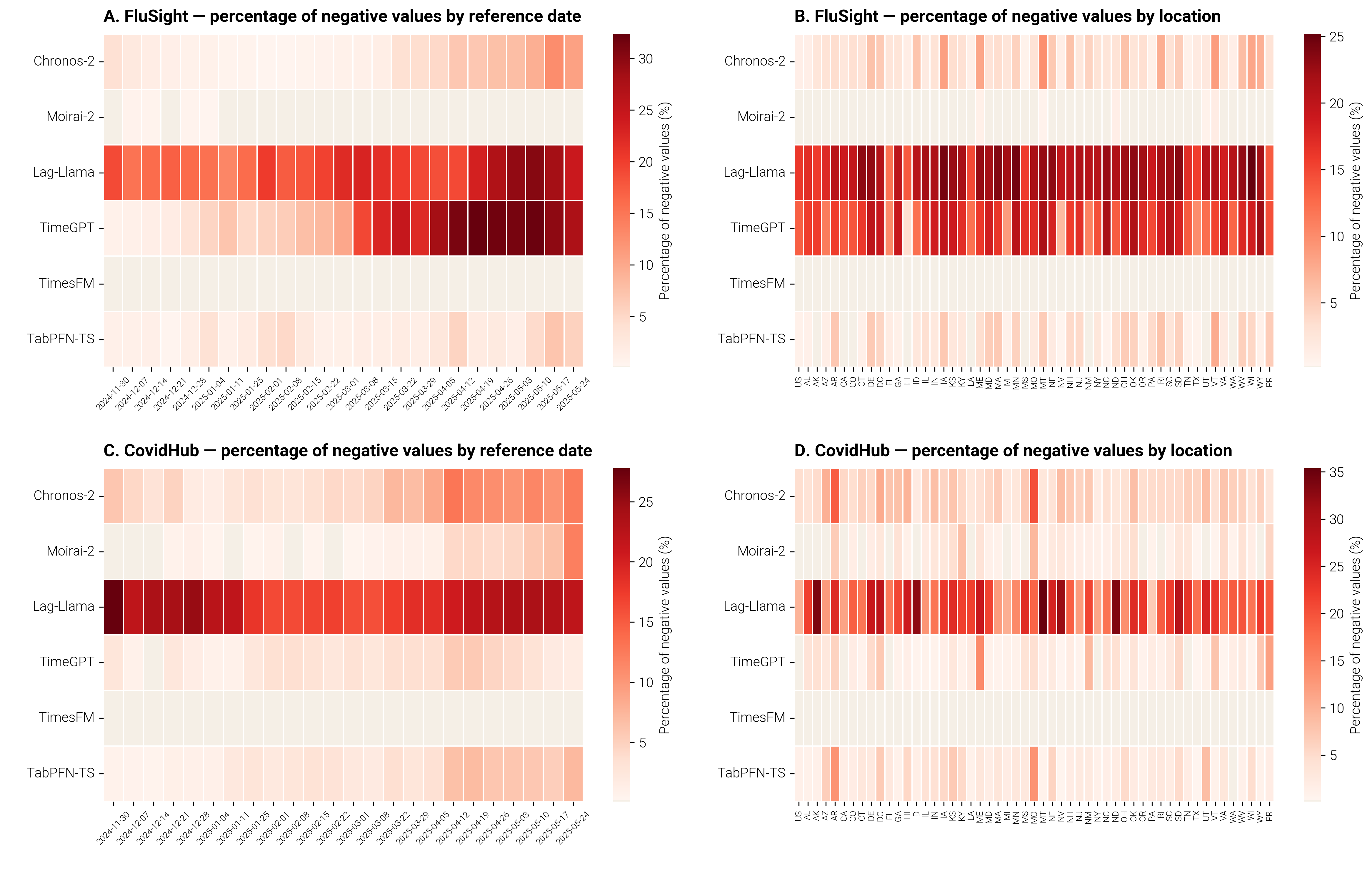

**Figure S9. Percentage of negative quantile values produced by each TSFM for FluSight and CovidHub, by reference date (A, C) and by location (B, D), season 2024/25.** For each model, colored cells indicate the percentage of quantile forecasts with negative predicted values, computed separately across all reference dates (A, C) and all locations (B, D), before values were clipped to zero. Cells shown in ivory correspond to a value of 0%. TSFM, time series foundation model.

Fig. S10 shows the percentage of quantile values requiring monotonicity correction through interpolation for each TSFM, broken down by reference date (A, C) and by location (B, D), during the 2025/26 season, for FluSight and CovidHub. Considering FluSight, only Chronos-2 and Moirai-2 required this correction, with Moirai-2 showing comparatively higher percentages, particularly for the earliest reference dates and across most locations; Lag-Llama, TimeGPT, TimesFM, and TabPFN-TS did not require quantile correction. Considering CovidHub, Chronos-2 shows a comparatively higher and more persistent percentage of quantile correction across most of the study period and across nearly all locations, while Moirai-2 required only occasional correction; as for FluSight, the remaining four TSFMs did not require quantile correction. Fig. S11 presents the same visualization for the 2024/25 season. Unlike in 2025/26, Chronos-2 required only marginal corrections for FluSight (last reference date), though it still did for CovidHub; conversely, Moirai-2 required only minimal corrections for CovidHub. As in 2025/26, all other TSFMs did not require any corrections.

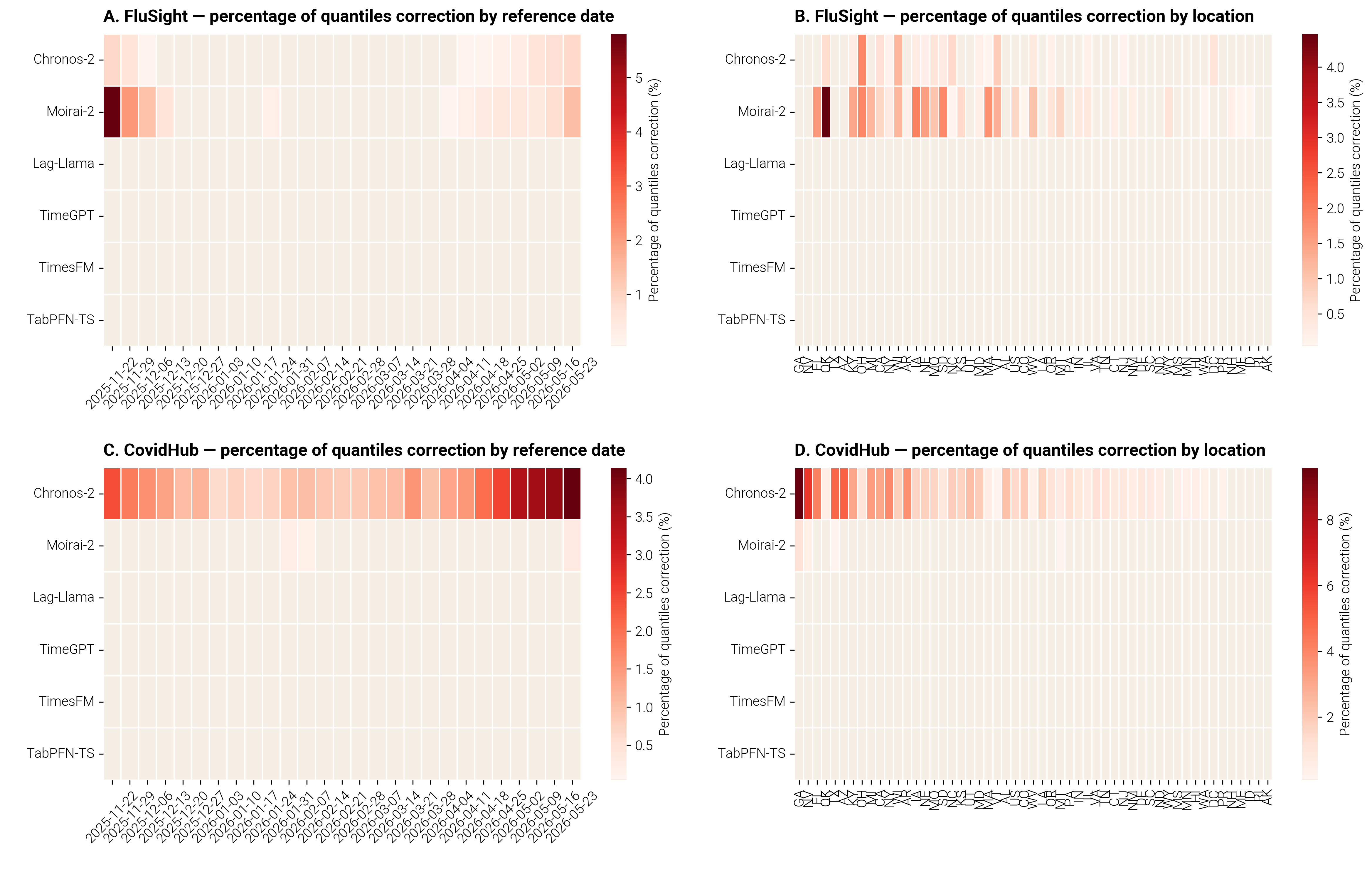

**Figure S10. Percentage of quantile levels requiring correction for each TSFM in FluSight and CovidHub, by reference date (A, C) and by location (B, D), season 2025/26.** For each model, colored cells indicate the percentage of quantile values violating monotonicity and corrected via linear interpolation, computed separately across all reference dates (A, C) and all locations (B, D). Cells shown in ivory correspond to a value of 0%. TSFM, time series foundation model.

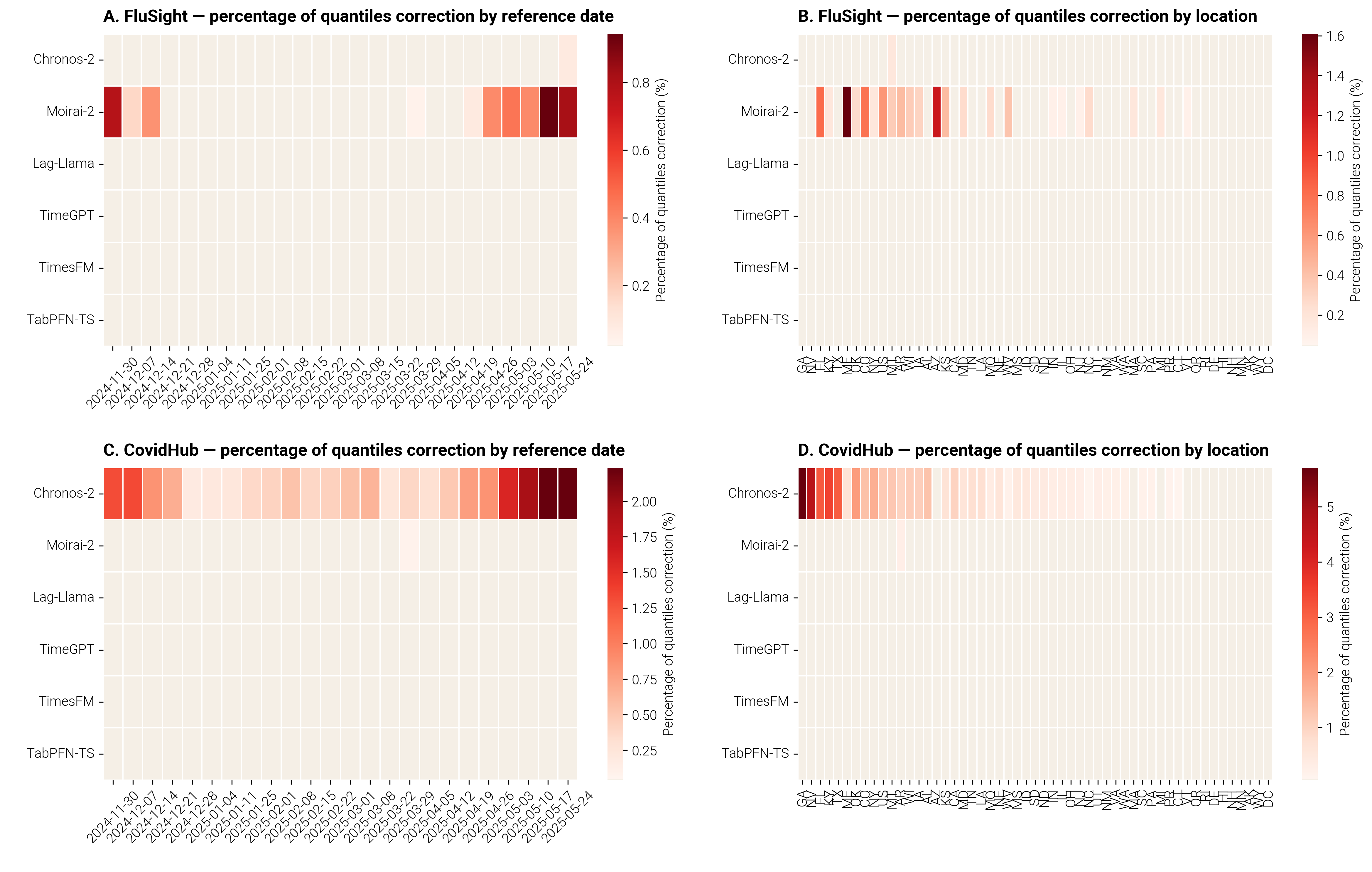
**Figure S11. Percentage of quantile levels requiring correction for each TSFM in FluSight and CovidHub, by reference date (A, C) and by location (B, D), season 2024/25.** For each model, colored cells indicate the percentage of quantile values violating monotonicity and corrected via linear interpolation, computed separately across all reference dates (A, C) and all locations (B, D). Cells shown in ivory correspond to a value of 0%. TSFM, time series foundation model.

**S5 Sensitivity analysis on trend classification**

Here, we present the sensitivity analysis on the obtained thresholds for trend classification, comparing with the official one reported by the CDC in Ref. [1]. The official thresholds are the following: stable weeks were defined as a change in weekly rate ≥ −0.3 to < 0.3 influenza-associated hospitalizations/100,000, or as weeks with an absolute difference of <10 hospital admissions. Increases were defined as a positive change in weekly rate ≥ 0.3 to < 1.7/100,000, and large increases as a positive change ≥ 1.7/100,000. Decreases were defined as a negative change in weekly rate > −1.7 to < −0.3/100,000, and large decreases as a negative change ≤ −1.7/100,000.

In Figure S12, we present the week-by-week classification on the U.S. national influenza epidemic dynamics for both 2024/25 and 2025/26, comparing the CDC official methodology and the adapted one. In 2024/25, the adapted method largely agreed with CDC official classifications: increase, large decrease, and large increase weeks were classified identically 100% of the time. Agreement was slightly lower for decrease weeks, where 80% were classified the same, while the remaining 20% were reclassified as large decrease. The largest discrepancy occurred for stable weeks, where the adapted method agreed with the official classification 76.7% of the time, while 13.3% were reclassified as increase and 10% as decrease. In 2025/26, agreement between the adapted method and CDC official classifications was even higher for most categories: decrease, increase, large decrease, and large increase weeks were all classified identically 100% of the time. As in 2024/25, the largest discrepancy occurred for stable weeks, where the adapted method agreed with the official classification 69.6% of the time, while 21.7% were reclassified as increase and 8.7% as decrease.

Considering all the U.S. locations and both seasons, the overall percentage of agreement was 89%. Agreement was highest for large decrease and large increase weeks, both classified identically 100% of the time, and remained high for stable weeks (86.5%) and increase weeks (97.3%, with the remaining 2.7% reclassified as large increase). Agreement was comparatively lower for decrease weeks, where 84.8% were classified the same and the remaining 15.2% were reclassified as large decrease.

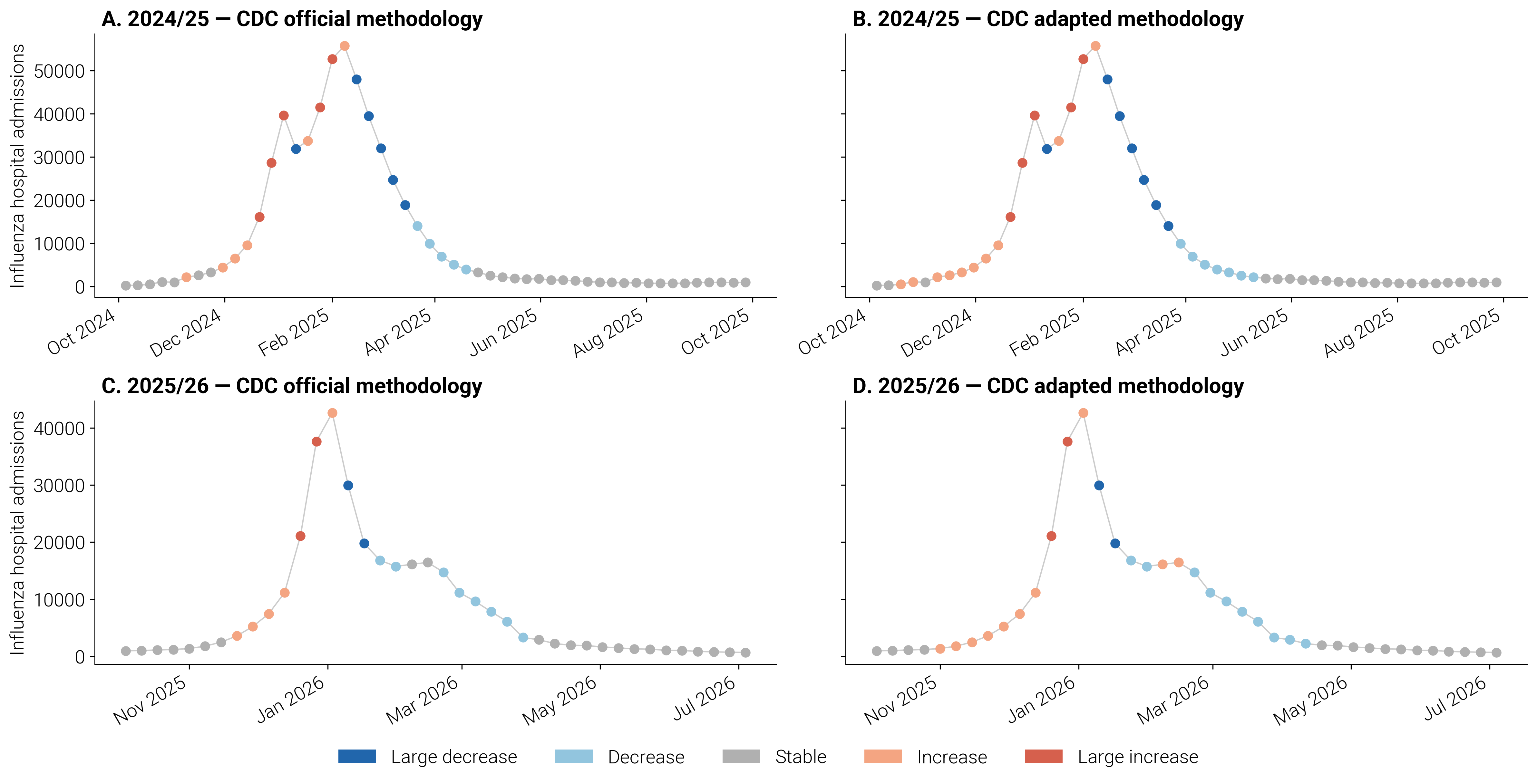
**Figure S12.** **Comparison of trend classifications derived from the CDC official and adapted methodologies for the U.S. national influenza epidemic curve.** Weekly influenza hospital admissions in the United States, colored by trend category (large decrease, decrease, stable, increase, large increase), for the 2024/25 (A, B) and 2025/26 (C, D) seasons. Panels A and C show trend classifications derived using the original CDC methodology thresholds [1]. Panels B and D show classifications derived using our adapted methodology, with thresholds calculated directly from NHSN hospital admissions data over a period from 2022/23 to 2025/26.
